# Outcomes following out-of-hours cholecystectomy: A systematic review and meta-analysis

**DOI:** 10.1101/2021.02.03.21251096

**Authors:** Sameer Bhat, Chris Varghese, William Xu, Ahmed W.H. Barazanchi, Bathiya Ratnayake, Gregory O’Grady, John A. Windsor, Cameron I. Wells

**Affiliations:** Department of Surgery, Faculty of Medical and Health Sciences, The University of Auckland, Auckland, New Zealand; Auckland Bioengineering Institute, The University of Auckland, Auckland, New Zealand

**Keywords:** Urgent cholecystectomy, post-operative complications, night, weekend, patient safety

## Abstract

**Background:** Cholecystectomy is one of the most commonly performed abdominal operations. Demands on acute operating theatre availability have led to out-of-hours (evenings, nights, or weekend) cholecystectomy being performed, although it is not known whether outcomes differ between out-of-hours and in-hours (daytime on weekdays) cholecystectomy.

**Objective:** This systematic review and meta-analysis aimed to compare outcomes following out-of-hours *versus* in-hours urgent cholecystectomy.

**Methods:** MEDLINE, EMBASE and Scopus databases were systematically searched from inception to December 2020 for studies comparing outcomes from out-of-hours and in-hours urgent cholecystectomy in adults. The outcomes of interest were rates of bile leakage, bile duct injury (BDI), overall post-operative complications, conversion to open cholecystectomy, specific intra- and post-operative complications, length of stay (LOS), readmission and mortality. Sensitivity analysis of adjusted multivariate results was also performed.

**Results:** In total, 194,135 urgent cholecystectomies (30,001 out-of-hours; 164,134 in-hours) from 11 studies were included. Most studies were of high (64%) or medium (18%) quality. There were no differences between out-of-hours and in-hours cholecystectomy for rates of bile leakage, BDI, overall post-operative complications, conversion to open cholecystectomy, operative duration, readmission, mortality, and post-operative LOS. Higher rates of post-operative sepsis (odds ratio (OR) 1.58, 95% CI: 1.04-2.41; p=0.03) and pneumonia (OR 1.55, 95% CI: 1.06-2.26; p=0.02) were observed following out-of-hours cholecystectomy on univariate meta-analysis but not on adjusted multivariate meta-analysis.

**Conclusions:** There was no increased risk or difference in specific complications associated with out-of-hours compared with in-hours urgent cholecystectomy.

## INTRODUCTION

Cholecystectomy is one of the most commonly performed abdominal operations and the gold standard treatment for acute benign gallbladder disease ^1,2^. In patients with acute cholecystitis, urgent cholecystectomy (i.e. non-elective, index cholecystectomy) is associated with fewer post-operative complications ^3^, improved resource utilization and timeliness of care compared with elective cholecystectomy in this acute setting ^4,5^. Increasing demands on acute operating theatre availability, as well as potential reductions in hospital length of stay (LOS) ^6,7^, have led to some centres performing urgent cholecystectomy out-of-hours (in the evenings, at night, or on weekends) ^8^.

However, it is not known whether out-of-hours and in-hours (during the daytime on weekdays) urgent cholecystectomy have different outcomes. Some studies suggest that out-of-hours cholecystectomy is associated with increased rates of conversion to open surgery and risk of complications ^9,10^. Yet several studies have argued that no such association exists, citing similar rates of conversion to open surgery, biliary complications and total LOS for out-of-hours and in-hours urgent cholecystectomy ^11–13^.

We hypothesized that out-of-hours cholecystectomy may more be likely to be performed by less experienced surgeons on more unwell patients during unsocial hours, and therefore would be associated with higher risk of intra-operative and post-operative complications. Therefore, this systematic review and meta-analysis aimed to compare outcomes following out-of-hours and in-hours urgent cholecystectomy.

## METHODS

This study was reported in accordance to the Preferred Reporting Items for Systematic Reviews and Meta-Analyses (PRISMA) and Meta-analysis of Observational Studies in Epidemiology (MOOSE) guidelines ^14,15^; PRISMA and MOOSE checklists are shown in **Supplementary Appendix 1**. The study protocol was prospectively registered on the PROSPERO registry (ID: CRD42021226127) prior to the database search.

### Search strategy

The MEDLINE (OVID), EMBASE (1980-2020), and Scopus databases were systematically searched from inception to December 16^th^, 2020. The following keywords and Medical Subject Headings (MeSH) were combined with the Boolean “AND” and “OR” operators and adapted for each database: “urgent”, “cholecystectomy”, “out-of-hour” and “day-time”. An example of the complete search string is outlined in **Supplementary Appendix 2**. The search was restricted to studies of adult populations (≥18 years old) published in English; there were no limitations on the geographic location or design of studies. Reference lists of included studies and potentially relevant reviews were also screened to ensure all relevant studies were captured.

### Study selection

All original research articles comparing intra-operative and post-operative outcomes in adults (≥18 years old) undergoing out-of-hours *versus (vs*.) in-hours urgent cholecystectomy for any acute benign gallbladder disease were eligible for inclusion. “Out-of-hours” cholecystectomy was defined using the author’s definition, however as a minimum requirement, out-of-hours cases must have commenced after 1600 hours on a weekday or at any time on a weekend. Cholecystectomy was considered urgent if it was performed during the patient’s index hospital admission (i.e. non-elective) with acute gallbladder disease.

All letters, commentaries, review articles, case reports (n <5 patients), conference abstracts, and studies with pediatric patients (≤18 years old) were excluded. Studies where the timing of cholecystectomy (time of day and/or day of the week) was not provided, or where direct comparisons between out-of-hours and in-hours groups were not made, were all excluded. Studies of elective cholecystectomy and non-operative management for acute benign gallbladder disease were also excluded.

Study titles and abstracts were independently screened for their eligibility by two authors. Full-texts were derived and independently reviewed by two authors. Any discrepancies were resolved through discussion amongst authors.

### Data extraction

Data from included studies were extracted into a pro forma spreadsheet by one author and validated for accuracy by a second independent author. Disagreements between authors were mediated by a senior author. Extracted data were characterised as follows: study characteristics; definitions and criteria; surgical service characteristics; pre-operative/patient demographic variables; planned operative approach; intra-operative variables and post-operative complication rates. The full list of extracted data is tabulated in **Table S1**. For included papers that reported a multivariate analysis, the adjusted odds ratio (OR), 95 percent (%) confidence interval (CI), and covariates that were adjusted for in each model, were extracted. Outcome definitions were also extracted and tabulated. Authors were contacted for missing data where possible.

### Quality assessment

Methodological appraisal of studies were conducted using the Newcastle-Ottawa Quality Assessment Scale (NOS) ^16^, and the Joanna Briggs Institute (JBI) Critical Appraisal Checklist for Cohort Studies ^17^. Studies with eight or more stars on the NOS were considered high-quality, seven stars were considered medium-quality and six or fewer stars were low-quality ^16^.

### Outcome measures

The outcomes of interest were rates of overall biliary complications, overall post-operative complications, conversion to open cholecystectomy, time from hospital admission to cholecystectomy, operative duration, intra-operative blood loss, intra-operative cholangiogram (IOC) use, total and post-operative LOS, specific post-operative complications, readmission, mortality, and whether the critical view of safety was obtained ^18^. Overall biliary complications were defined as the pooled rate of bile leak and bile duct injury (BDI); however, bile leakage and BDI were analyzed separately where possible. Overall post-operative complications were defined as any complication or morbidity (i.e. at least one grade 1 Clavien-Dindo (CD) complication ^19^), including biliary complications, occurring within the index hospital admission or up to 90 days post-operatively, although individual study definitions were extracted when reported. Conversion to open cholecystectomy was defined as the proportion of cholecystectomies planned as laparoscopic which were converted to an open procedure intra-operatively. Specific post-operative complications comprised wound infection, intra-abdominal abscess, bleeding, sepsis, and pneumonia. Outcomes were defined according to definitions provided by each author, and these were also extracted and tabulated where possible.

### Statistical analysis

Unadjusted continuous and categorical data from univariate analyses of each study were reported as the mean ± standard deviation (SD) and frequency (%), respectively. Continuous data that were originally reported as the median and range or interquartile range were converted to the mean (via the methods of Luo *et al*. ^20^) and SD (via the methods of Wan *et al*. ^21^). Continuity corrections were applied to dichotomous (categorical) outcomes with zero events, by adding one to both the numerator and denominator of each study arm ^22^. Pooled summary estimates for the outcomes of interest were reported as the mean difference (MD) or OR for continuous and dichotomous outcomes, respectively ^23^, with their 95% CI. A random-effects model with the DerSimonian-Laird tau estimator for between study variance was used ^24^. Forest plots were used to visualise meta-analysis results. Heterogeneity between the included studies were measured using the I^2^ statistic ^25^. An I^2^ value of 0-40% was considered not significantly heterogeneous, 30-60%: moderate, 50-90%: substantial and 75-100% considerable heterogeneity ^26^. Publication bias was assessed via visual inspection of funnel plots for asymmetry.

Subgroup analyses of evenings and night-time *vs*. daytime urgent cholecystectomy (excluding studies where only the day of the week were reported), weekend *vs*. weekday urgent cholecystectomy (excluding studies where day of the week were not reported) and urgent cholecystectomy planned laparoscopically (excluding studies with planned open cholecystectomy) was performed for the outcomes of interest. Authors were individually contacted for their original data in studies where planned open and laparoscopic cholecystectomy outcomes could not be separated for the purposes of subgroup analysis ^12,27–29^. A sensitivity analysis including only studies with multivariate regression analyses for any of the outcomes was also performed. The 95% CI from the multivariate analysis of each study was converted to the standard error (SE) ^26^. OR and SE for each study were pooled using the generic inverse-variance meta-analysis method and reported as the pooled OR with 95% CI for each outcome. P values <0.05 were considered statistically significant. All statistical analyses were conducted using R (Version 4.0.2; R Foundation for Statistical Computing, Vienna, Austria) ^30^.

## RESULTS

### Search results

The database search found 6,965 results, 11 of which were included in the quantitative meta-analysis ^6,9–13,27–29,31,32^ (refer PRISMA flow diagram, **Supplementary Appendix 3**). Studies were mostly excluded because they either did not perform direct comparisons between out-of-hours and in-hours cholecystectomy (n = 7), the timing of cholecystectomy was not provided (n = 4) or related to conference abstracts only (n = 4).

### Study characteristics

The study design and characteristics of each study are shown in **Table 1**. All were retrospective cohort studies conducted between 2009 and 2020. Most involved a single centre (n = 6), and the majority of studies were conducted in the United States (n = 6) ^6,9,10,13,27,28^. The selection and diagnostic criteria and out-of-hours definitions of each study are provided in **Table S2**. Definitions relating to out-of-hours urgent cholecystectomy varied between studies, but there were two studies that used 0700 and 1900 as cut-offs ^10,13^. Three studies considered only urgent cholecystectomy performed on weekends as out-of-hours (i.e. time of day were not provided) ^27–29^.

**Table 1.** Study design and characteristics of included studies.

| Author | Year | Country | Number of centres | Study period | Study design |
| --- | --- | --- | --- | --- | --- |
| Geraedts <sup>11</sup> | 2018 | the Netherlands | 1 | 1<sup>st</sup> January 2014 – 31<sup>st</sup> December 2016 | Retrospective observational cohort study |
| Gustafsson <sup>12</sup> | 2020 | Sweden | Population based <sup>a</sup> | 2006 – 2017 | Retrospective observational cohort study |
| Hoehn <sup>27</sup> | 2018 | United States | 118 <sup>b</sup> | 2009 – 2013 | Retrospective observational cohort study |
| Li <sup>31</sup> | 2009 | China | 1 | January 2006 – December 2007 | Retrospective observational cohort study |
| Phatak <sup>9</sup> | 2014 | United States | 1 | October 2010 – May 2011 | Retrospective observational cohort study |
| Rothman <sup>29</sup> | 2016 | Denmark | Not specified <sup>c</sup> | 2006 – 2011 | Retrospective observational cohort study |
| Siada <sup>6</sup> | 2017 | United States | 1 | October 2011 – June 2015 | Retrospective observational cohort study |
| Tseng <sup>13</sup> | 2019 | United States | 1 | April 2007 – February 2015 | Retrospective observational cohort study |
| Wu <sup>10</sup> | 2014 | United States | 2 | 1<sup>st</sup> January 2011 – 1<sup>st</sup> August | Retrospective observational cohort study |
| Yoshioka <sup>32</sup> | 2019 | Japan | 1 | January 2005 – December 2017 | Retrospective observational cohort study |
| Zapf <sup>28</sup> | 2015 | United States | Not specified <sup>d</sup> | 2007 – 2010 | Retrospective observational cohort study |

<sup>a</sup> Data were obtained from the Swedish National Register for Gallstone Surgery and Endoscopic Retrograde Cholangiopancreatography (GallRiks).
<sup>b</sup> Data obtained from the University HealthSystem Consortium, comprising 118 academic medical centres and 298 affiliated hospitals.
<sup>c</sup> Patients were registered within the Danish Cholecystectomy Database.
<sup>d</sup> Patients registered with the Healthcare Cost and Utilization Project Florida State Inpatient Database were analyzed.

### Patient and procedure characteristics

Urgent cholecystectomies were performed in a total of 194,135 patients, among which 30,001 (15.5%) patients underwent out-of-hours operations and 164,134 (84.5%) patients underwent in-hours operations. Of the 194,135 included urgent cholecystectomies, 179,837 were planned laparoscopically and 2,367 were planned as open procedures; Zapf *et al*. ^28^ did not specify their operative approach. Patient demographic descriptors and pre-operative variables for each study are summarized in **Table 2**.

**Table 2.** Patient demographic and pre-operative variables in included studies

| Author | Sample size, n | | Sex (M:F), n | | Mean ± SD (IQR), years | | BMI, kg/m<sup>2</sup> | | ASA Classification, n I/II/III/IV/V | | Diagnosis, n AC/CC/GC/GP/BC | | Tokyo severity grade for AC<sup>14</sup>, n I/II/III | |
| --- | --- | --- | --- | --- | --- | --- | --- | --- | --- | --- | --- | --- | --- | --- |
|  | OOH | IH | OOH | IH | OOH | IH | OOH | IH | OOH | IH | OOH | IH | OOH | IH |
| Geraedts<sup>11</sup> | 145 | 67 | 60:85 | 34:33 | 59.5 ± 13.3 | 58.1 ± 13.8 | | | | | 111/0/0/0/23 | 53/0/0/0/5 | | |
| Gustafsson<sup>12</sup> | 2,710 | 8,443 | 1,333:1,375 | 4,071:4,368 |  |  |  |  | 948/1,345/375/40/2 | 3,170/4,142/1,077/52/2 | 2,710/0/0/0/0 | 8,443/0/0/0/0 |  |  |
| Hoehn<sup>27</sup> | | | | | 47.4 ± 21.5 | 49.3 ± 22.2 | | | | | | | | |
| Li<sup>31</sup> | 19 | 64 | 12:7 | 27:37 | 63.8 ± 15.6 | 65.7 ± 16.6 | | | | | 19/0/0/0/0 | 64/0/0/0/0 | | |
| Phatak<sup>9</sup> | 108 | 248 | 18:90 | 49:199 | 34.8 ± 11.6 | 38.4 ± 14.0 | 29.4 ± 5.0 | 30.0 ± 5.7 | | | 80/10/0/13/0 | 165/32/0/35/0 | 23/80/5 | 59/166/23 |
| Rothman<sup>29</sup> | 1,343 | 27,416 | 514:829 | 7,753:19,663 |  |  |  |  | 671/526/123/19/4 | 15,278/10,504/1,539/75/20 | 951/163/0/0/0 | 4,588/7,670/0/0/0 |  |  |
| Siada<sup>6</sup> | 219 | 647 | 62:157 | 183:464 | 42 ± 16 | 41 ± 16 | 32 ± 7 | 32 ± 7 | | | 219/0/0/0/0 | 647/0/0/0/0 | | |
| Tseng<sup>13</sup> | 576 | 4,628 | 110:466 | 817:3,811 | 36.4 ± 12.6 | 36.7 ± 13.3 | 30.6 ± 6.1 | 31.1 ± 6.2 | 60/139/30/1/0 | 349/1,297/80/5/0 | | | | |
| Wu<sup>10</sup> | 232 | 908 | 55:177 | 275:633 | 40.5 ± 2.7 | 40.9 ± 1.3 | | | 44/160/27/1/0 | 91/636/174/6/0 | 125/132/40/0/0 | 579/548/136/0/0 | | |
| Yoshioka<sup>32</sup> | 101 | 270 | 69:32 | 173:97 | 66 ± 14 | 64 ± 15 | 24.7 ± 3.7 | 24.8 ± 4.3 | 18/67/16/0/0 | 49/192/29/0/0 | 101/0/0/0/0 | 270/0/0/0/0 | 32/63/6 | 118/144/8 |
| Zapf<sup>28</sup> | 2,247 | 9,664 |  |  |  |  |  |  |  |  |  |  |  |  |

AC, acute cholecystitis; ASA, American Society of Anesthesiologists; BC, biliary colic; BMI, body mass index; CC, chronic cholecystitis; F, female; GC, gangrenous cholecystitis; GP, gallstone pancreatitis; IH, in-hours; IQR, interquartile range; M, male; n, number; OOH, out-of-hours.
Empty cells relate to data points that were not reported.

### Surgical service

The structure and characteristics relating to each surgical service are tabulated in **Table S3**. In four studies, data were obtained from large registries or population-based databases and hence the structure and/or characteristics of individual centres or institutions could not be ascertained ^12,27–29^. Of the remaining studies, three had an acute surgical unit (ASU) ^6,9,13^, with 24/7 access to this service for patients in two of these studies ^6,9^. Out-of-hours urgent cholecystectomy was performed by surgeons with diverse levels of training and experience (see **Table S3** for details of surgeons performing out-of-hours cholecystectomy).

### Quality assessment

The NOS scores for all included studies are illustrated in **Table S4**. Most studies were judged to be of either high (64%, 7/11) or medium (18%, 2/11) quality. Only two studies were judged as low-quality, principally due to the lack of any adjusted analysis to control for potentially confounding covariates ^6,13^. Results of the JBI Critical Appraisal Checklist for Cohort Studies are provided in **Table S5**. Completeness of follow up could not be evaluated due to the retrospective design of included studies and outcomes were not reliably recorded in three studies ^9,27,31^.

#### Outcome measures

Outcome definitions varied in each study (refer **Table S6** for outcome definitions in each study).

### Biliary complications

Overall biliary complication rates following urgent cholecystectomy were reported in two studies, comprising 5,560 operations (684 out-of-hours; 4,876 in-hours) ^9,13^. There were no differences in the rate of biliary complications between out-of-hours *vs*. in-hours urgent cholecystectomy on univariate meta-analysis (OR 1.11, 95% CI: 0.41 to 2.99; p=0.83; **Figure 1A**). Multivariate analyses were not conducted in these studies. On subgroup analysis, there were no differences in either the rate of bile leakage (OR 1.00, 95% CI: 0.58 to 1.74; p=0.99; **Figure 1B**) ^9,11,32^ or BDI (OR 0.98, 95% CI: 0.57 to 1.70; p=0.95; **Figure 1C**) ^6,9,12^. No significant heterogeneity was detected (I^2^ = 0%).

**Figure 1:**
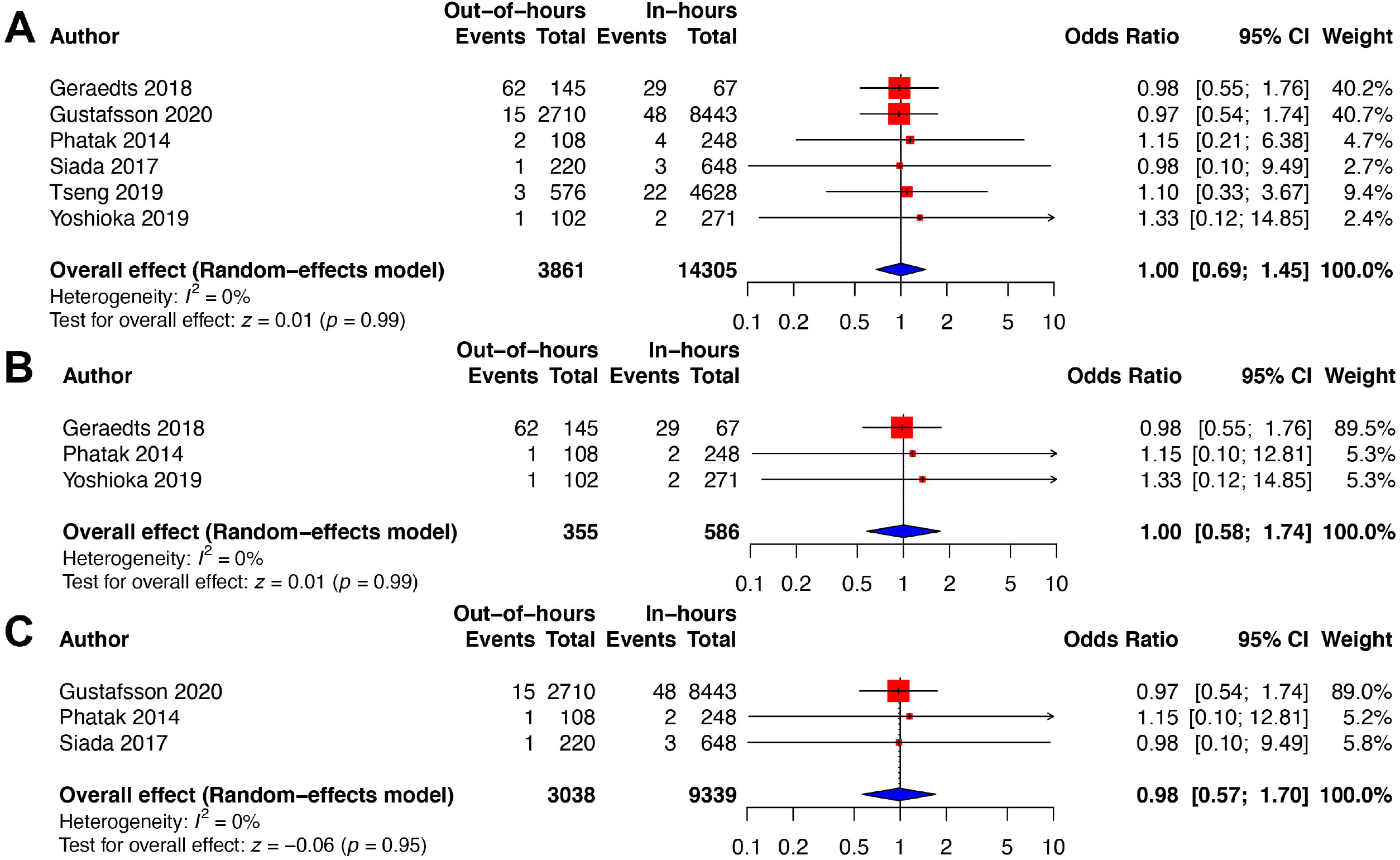
Forest plots of *(A)* overall biliary complications, *(B)* bile leakage, and *(C)* bile duct injury.

### Overall post-operative complications

Overall complication rates following urgent cholecystectomy were reported in five studies, comprising 13,886 operations (3,370 out-of-hours; 13,886 in-hours) ^6,9,10,12,32^. There were no differences in overall complication rates following out-of-hours *vs*. in-hours urgent cholecystectomy (OR 1.13, 95% CI: 0.93 to 1.36; p=0.21; **Figure 2**). Multivariate analyses were performed on four studies ^9–12^ and results did not differ on adjusted analysis (OR 1.04, 95% CI: 0.90 to 1.19; p=0.59). No significant heterogeneity was detected in pooled univariate or multivariate analyses (I^2^ = 10% and 0%, respectively).

**Figure 2:**
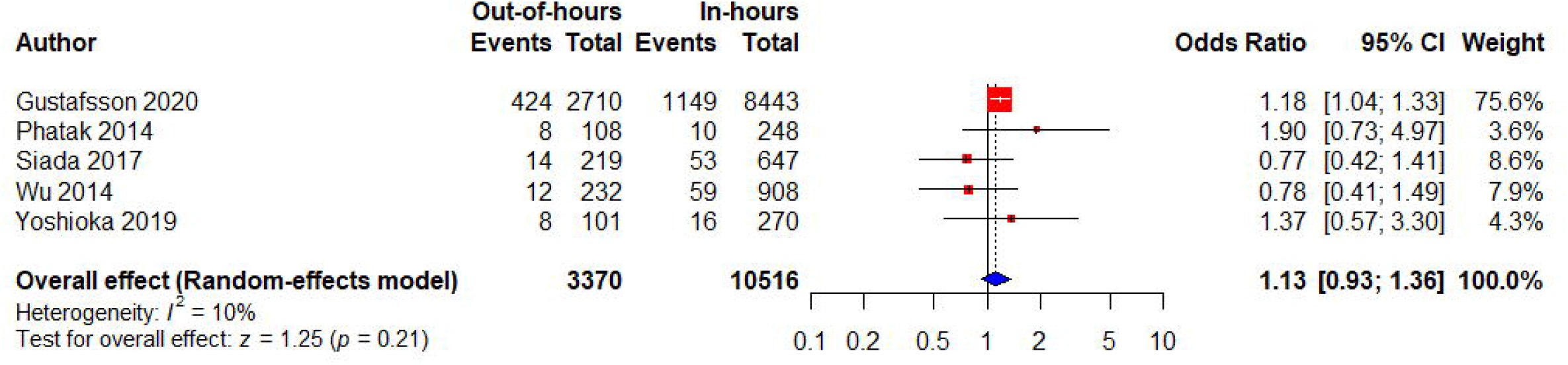
Forest plot of overall post-operative complications on univariate meta-analysis.

### Conversion to open cholecystectomy

Rates of conversion to open cholecystectomy were reported in eight studies, comprising 48,061 operations (5,434 out-of-hours; 42,627 in-hours) ^6,9–13,29,32^. No significant differences in rates of conversion to open cholecystectomy were found on univariate meta-analysis (OR 1.22, 95% CI: 0.74 to 2.02; p=0.44; **Figure 3**). Results of the multivariate analysis also did not differ (OR 1.19, 95% CI: 0.89 to 1.60; p=0.24) ^10,29,31^. There was considerable heterogeneity among studies on univariate (I^2^ = 94%) but less on multivariate (I^2^ = 22%) meta-analyses.

**Figure 3:**
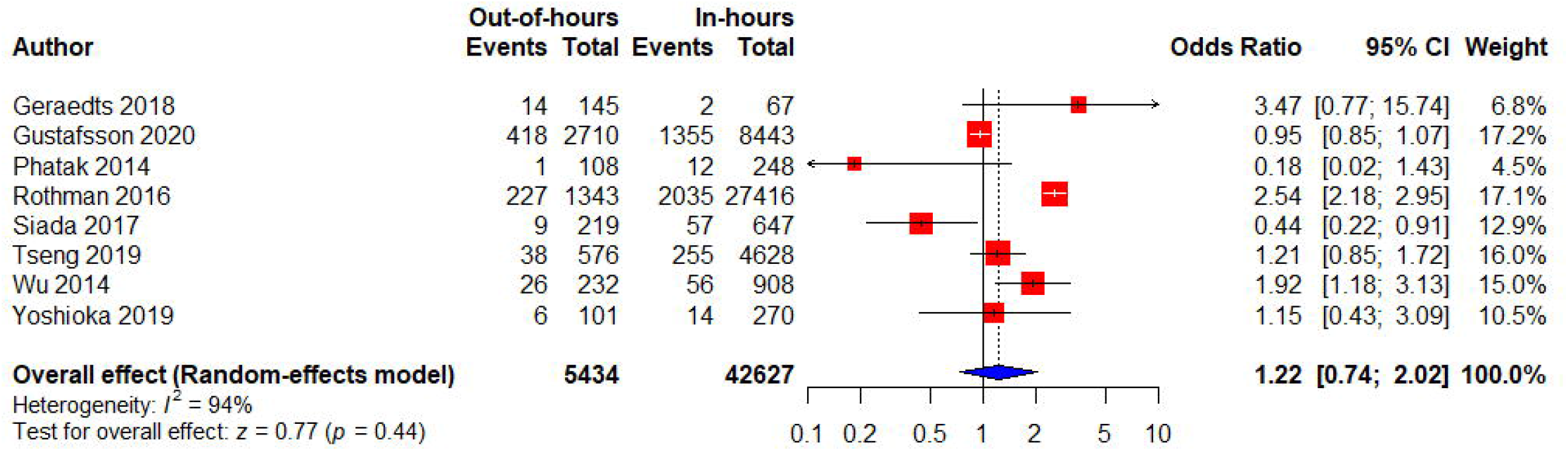
Forest plot of conversion to open cholecystectomy on univariate meta-analysis.

The meta-analyses of remaining outcomes are reported in **Supplementary Appendix 4**. Out-of-hours urgent cholecystectomy was associated with higher intra-operative blood loss (MD +1.49 mL, 95% CI: 0.21 to 2.78 mL; p=0.02) and lower rates of IOC (OR 0.68, 95% CI: 0.49 to 0.93; p=0.02), compared to in-hours cholecystectomy. Higher rates of post-operative sepsis (OR 1.58, 95% CI: 1.04 to 2.41; p=0.03) and pneumonia (OR 1.55, 95% CI: 1.06 to 2.26; p=0.02) were also observed on univariate meta-analysis, but not after the adjusted multivariate meta-analysis (OR 0.81, 95% CI: 0.41 to 1.58; p>0.05 and OR 1.10, 95% CI: 0.63 to 1.94; p>0.05, respectively). No differences were seen in the time from admission to cholecystectomy, operative duration, post-operative and total LOS, rates of wound infection, intra-abdominal abscesses, post-operative bleeding, readmission, and mortality. Geraedts *et al*. found no difference in the proportion of out-of-hours *vs*. in-hours operations in which the critical view of safety was obtained (86.2% *vs*. 91.0%; p=0.43) ^11^.

### Subgroup analysis

Subgroup analyses for evenings and night-time *vs*. daytime, weekend *vs*. weekday, and laparoscopic-only urgent cholecystectomy are tabulated in **Supplementary Appendix 5**. Time from hospital admission to cholecystectomy was less in evenings and night-time *vs*. daytime (MD -0.38 days, 95% CI: -0.48 to -0.27 days; p<0.0001; I^2^ = 0%) but not weekend *vs*. weekday (MD 0.70 days, 95% CI: 0.68 to 0.72 days; p<0.0001) urgent cholecystectomy. Other outcomes did not differ from the overall analyses.

### Sensitivity analysis

The covariates adjusted for in multivariate models for each study are reported in **Table S7**. Studies adjusted for a range of demographic and pre-operative variables; multivariate analyses were not performed for outcomes other than rates of overall post-operative complications, conversion to open cholecystectomy, and the specific post-operative complications of sepsis and pneumonia.

### Publication bias

Quantitative assessment via Egger’s regression test was not possible due to <10 studies being available for pooling. Symmetrical funnel plots were seen in all outcomes except for time from admission to cholecystectomy (refer **Supplementary Appendix 6** for funnel plots of the outcomes of interest).

## DISCUSSION

This systematic review and meta-analysis compared outcomes following out-of-hours *vs*. in-hours urgent cholecystectomy in adults with acute benign gallbladder disease. The meta-analysis found no difference in the rates of biliary complications, such as bile leak and BDI, overall post-operative complications, and conversion to open cholecystectomy. These results were also confirmed on subgroup analyses. Longer times from admission to cholecystectomy were observed during weekends compared to weekdays and shorter times during evening and night-time compared to daytime.

The rising demand for elective surgery, limited daytime operating theatre availability, and decreased staffing capacity, has meant urgent and semi-urgent operations such as cholecystectomy are performed out-of-hours in some centres ^7,8^. Importantly, this study found that this approach is not associated with an increase in complications across a large sample of 194,135 procedures. An increased risk of post-operative sepsis and pneumonia was observed in the out-of-hours cholecystectomy cohort on univariate meta-analysis, but not after adjusting for confounding patient and hospital factors on multivariate analysis. The higher risk before controlling for patient factors suggests out-of-hours operations are performed on patients with more severe disease. In addition, hospital factors such as reduced staff numbers out-of-hours may result in reduced use of post-operative ward-based protocols such as early patient ambulation, which have been shown to decrease post-operative pneumonia risk ^33^. Similarly, reduced ward staff numbers out-of-hours could also contribute to the higher rates of sepsis among patients undergoing out-of-hours cholecystectomy, because of delays in diagnosis ^34^. The impact of these systemic factors requires further investigation in the context of out-of-hours cholecystectomy.

In framing our original hypothesis for this study, we considered evidence showing that fatigue and sleep deprivation contribute to suboptimal surgical performance and outcomes during out-of-hours procedures ^35,36^. Sleep deprivation is common among acute care surgeons; nearly two-thirds were acutely or chronically sleep deprived in a recent prospective study ^37^. Acute sleep deprivation may result in impaired attention, concentration, cognitive processing speed and response times and a lower threshold for stress ^36,38^, and can contribute to medical errors and poorer patient outcomes ^39^. Despite these known associations, it was notable that this meta-analysis did not demonstrate any differences in outcomes between out-of-hours and in-hours cholecystectomy, demonstrating that surgical services generally maintain high-quality outcomes for this common procedure. It is also unlikely that many out-of-hours cholecystectomies were performed between the hours of 12 AM and 6 AM, when surgical services usually revert to only critical activities ^40^. In addition, our results should not be generalised to other out-of-hours procedures, and further research should continue to address the important impacts of fatigue and sleep deprivation on surgeons and surgical outcomes ^37,38,41^.

One area of difference in practice noted on the present meta-analysis was lower rates of IOC in the out-of-hours cholecystectomy patient cohort. A recent meta-analysis of over 2 million patients showed that routine IOC use decreases the risk of BDI (0.36% *vs*. 0.53% for selective IOC) ^42^, but this trend was not apparent in out-of-hours cases showing lower IOC rates in our relatively smaller study (n = 194,135). The lower rates of IOC in the out-of-hours cohort might be explained by surgeons being less experienced, radiographers not being as available for imaging, or technical issues with more severe acute cholecystitis.

The meta-analysis did not find any difference in post-operative mortality rates following out-of-hours and in-hours cholecystectomy. This contrasts the findings of a recent meta-analysis which found higher rates of in-hospital and 30-day post-operative mortality following night-time/after-hours compared to daytime surgery in general ^43^. However, Cortegiani *et al*. included a much broader and more complex range of procedures in their analysis, comprising major urological and vascular, orthopaedic, trauma, cardiac and transplant surgery, and with both elective and non-elective surgeries included in the analysis. The primary outcome of mortality utilized by Cortegiani *et al*. may not accurately reflect differences in outcomes of specific operations. Mortality is a relatively rare outcome in cholecystectomy ^44^ and procedure-specific outcomes such as conversion, bile leak, and BDI, which impact on morbidity and long-term quality of life, are more typical quality indicators ^45^.

Interestingly, a subgroup analysis found faster time from hospital admission to cholecystectomy at evenings and night-time than during daytime and slower times on weekends compared to weekdays, although the total hospital LOS were similar. The shorter times observed at night may result because patients had more severe acute cholecystitis and needed more urgent cholecystectomy. The observed delay to urgent cholecystectomy on weekends may reflect lower staff availability and operating theatre capacity on weekends ^46^.

The present review identified two studies in which patients had 24/7 access to surgical care and services through an ASU. The availability of an ASU has been shown to increase daytime surgical capacity and acute operating theatre availability ^7,47^. Lim *et al*. found that night-time cholecystectomies decreased from 26.1% to 7.9% following ASU establishment (p<0.0001) ^48^, and von Conrady *et al*., found the proportion of emergency procedures performed overnight to decrease from 27.7% to 14.4% with the introduction of an ASU ^7^. In the case of urgent cholecystectomy, one retrospective study failed to show a significant decrease in the proportion of out-of-hours operations after introduction of an ASU (20 to 16%, p=0.34) ^47^. While the impact of an ASU model could not be assessed in this meta-analysis, its effect on the outcomes of out-of-hours cholecystectomy is of interest and should be assessed in future studies.

This review has several limitations. The primary aim of this study was to compare intra- and post-operative outcomes following out-of-hours and in-hours cholecystectomy through meta-analysis. However, ‘out-of-hours’ was variably defined, and this review was unable to differentiate between evening and night-time (e.g. 12 AM to 6 AM) urgent cholecystectomy within the out-of-hours cohort. Future studies should consider specifically assessing outcomes following day-time *vs*. evening *vs*. night-time urgent cholecystectomy, as well as weekdays *vs*. weekends. Furthermore, clinical outcomes were variably defined and reported, which limited the ability to pool studies. Inconsistency and heterogeneity in reporting of cholecystectomy outcomes was recognized as a significant issue in a recent systematic review ^44^. Standardized outcome reporting should hence be the focus of future studies. In this study, a random effects meta-analysis model was used to mitigate the impact of this heterogeneity. The retrospective design of the included studies presents a selection bias with patients with more severe acute cholecystitis potentially being more likely to undergo urgent out-of-hours cholecystectomy. However, a sensitivity analysis of adjusted analyses was performed to attempt to account for this potential bias. Due to limitations in data reporting, the present analysis was unable to assess “bail out” options for difficult gallbladders other than rates of conversion to open surgery; subtotal cholecystectomy ^49^ and other approaches to difficult gallbladders should be reported by future studies.

In conclusion this systematic review and meta-analysis has not shown an increased rate of complications or conversion to open surgery for out-of-hours compared with in-hours urgent cholecystectomy. While out-of-hours cholecystectomy appears to be a safe practice that can be generally supported, night-time cholecystectomy (e.g. 12 AM to 6 AM) was not specifically evaluated and is likely to be uncommonly practised. The decision to perform routine out-of-hours cholecystectomy at an individual hospital level must ultimately depend on the sufficiency of local resources, ensuring compliance with safe surgical working hours, and the adequacy of senior surgical oversight.

## Data Availability

This study does not contain any primary data.

## LIST OF SUPPLEMENTARY APPENDICES

**Supplementary Appendix 1:**

PRISMA and MOOSA Checklists.

**Supplementary Appendix 2:**

Search string for the MEDLINE (OVID) database.

**Table S1:**

List of extracted data from included studies.

**Supplementary Appendix 3:**

PRISMA flow diagram outlining the selection process for included studies.

**Table S2:**

Selection criteria and out-of-hours definitions in included studies.

**Table S3:**

Structure and characteristics of each surgical service.

**Table S4:**

Methodological quality assessment of the included studies using the Newcastle-Ottawa Quality Assessment Scale.

**Table S5:**

Quality appraisal of included studies using the JBI Critical Appraisal Checklist for Cohort Studies.

**Table S6:**

Outcome definitions provided by each study.

**Supplementary Appendix 4:**

Meta-analyses and forest plots of outcomes of interest other than biliary complications, overall post-operative complications and conversion to open cholecystectomy following out-of-hours and in-hours urgent cholecystectomy.

**Supplementary Appendix 5:**

Subgroup analyses of evenings and night-time *vs*. daytime, weekend *vs*. weekday and laparoscopic-only urgent cholecystectomy.

**Table S7:**

Covariates included within the multivariate analysis for each study.

**Supplementary Appendix 6:**

Funnel plots for the outcomes of interest.

