## Supplementary Appendix for "Outcomes following out-of-hours cholecystectomy: A systematic review and meta-analysis"

#### SUPPLEMENTARY APPENDIX 1: PRISMA and MOOSE Checklists

| Section/topic | # | Checklist item | Reported on page # |
|---|---|---|---|
| **TITLE** | | | |
| Title | 1 | Identify the report as a systematic review, meta-analysis, or both. | 1 (Title Page) |
| **ABSTRACT** | | | |
| Structured summary | 2 | Provide a structured summary including, as applicable: background; objectives; data sources; study eligibility criteria, participants, and interventions; study appraisal and synthesis methods; results; limitations; conclusions and implications of key findings; systematic review registration number. | 2-3 |
| **INTRODUCTION** | | | |
| Rationale | 3 | Describe the rationale for the review in the context of what is already known. | 4 |
| Objectives | 4 | Provide an explicit statement of questions being addressed with reference to participants, interventions, comparisons, outcomes, and study design (PICOS). | 4 |
| **METHODS** | | | |
| Protocol and registration | 5 | Indicate if a review protocol exists, if and where it can be accessed (e.g., Web address), and, if available, provide registration information including registration number. | 5 |
| Eligibility criteria | 6 | Specify study characteristics (e.g., PICOS, length of follow-up) and report characteristics (e.g., years considered, language, publication status) used as criteria for eligibility, giving rationale. | 5-6 |
| Information sources | 7 | Describe all information sources (e.g., databases with dates of coverage, contact with study authors to identify additional studies) in the search and date last searched. | 5 |
| Search | 8 | Present full electronic search strategy for at least one database, including any limits used, such that it could be repeated. | 5, Appendix S2 |
| Study selection | 9 | State the process for selecting studies (i.e., screening, eligibility, included in systematic review, and, if applicable, included in the meta-analysis). | 5-6 |
| Data collection process | 10 | Describe method of data extraction from reports (e.g., piloted forms, independently, in duplicate) and any processes for obtaining and confirming data from investigators. | 6 |

| | | | |
|---|---|---|---|
| Data items | 11 | List and define all variables for which data were sought (e.g., PICOS, funding sources) and any assumptions and simplifications made. | 6-7 |
| Risk of bias in individual studies | 12 | Describe methods used for assessing risk of bias of individual studies (including specification of whether this was done at the study or outcome level), and how this information is to be used in any data synthesis. | 7 |
| Summary measures | 13 | State the principal summary measures (e.g., risk ratio, difference in means). | 8-9 |
| Synthesis of results | 14 | Describe the methods of handling data and combining results of studies, if done, including measures of consistency (e.g., $I^2$) for each meta-analysis. | 8-9 |
| **Section/topic** | **#** | **Checklist item** | **Reported on page #** |
| Risk of bias across studies | 15 | Specify any assessment of risk of bias that may affect the cumulative evidence (e.g., publication bias, selective reporting within studies). | 8-9 |
| Additional analyses | 16 | Describe methods of additional analyses (e.g., sensitivity or subgroup analyses, meta-regression), if done, indicating which were pre-specified. | 8-9 |
| **RESULTS** | | | |
| Study selection | 17 | Give numbers of studies screened, assessed for eligibility, and included in the review, with reasons for exclusions at each stage, ideally with a flow diagram. | 10, Appendix S3 |
| Study characteristics | 18 | For each study, present characteristics for which data were extracted (e.g., study size, PICOS, follow-up period) and provide the citations. | 10-11, Tables 1-2, S2 and S5 |
| Risk of bias within studies | 19 | Present data on risk of bias of each study and, if available, any outcome level assessment (see item 12). | 10, Table S3 and S4 |
| Results of individual studies | 20 | For all outcomes considered (benefits or harms), present, for each study: (a) simple summary data for each intervention group (b) effect estimates and confidence intervals, ideally with a forest plot. | 11-13 |
| Synthesis of results | 21 | Present results of each meta-analysis done, including confidence intervals and measures of consistency. | 11-13, Figures 1-3, Appendix S4 |
| Risk of bias across studies | 22 | Present results of any assessment of risk of bias across studies (see Item 15). | 14, Appendix |

| | | | |
|---|---|---|---|
| | | | S6 |
| Additional analysis | 23 | Give results of additional analyses, if done (e.g., sensitivity or subgroup analyses, meta-regression [see Item 16]). | 13, Appendix S5 and Table S7 |
| **DISCUSSION** | | | |
| Summary of evidence | 24 | Summarize the main findings including the strength of evidence for each main outcome; consider their relevance to key groups (e.g., healthcare providers, users, and policy makers). | 15-18 |
| Limitations | 25 | Discuss limitations at study and outcome level (e.g., risk of bias), and at review-level (e.g., incomplete retrieval of identified research, reporting bias). | 18 |
| Conclusions | 26 | Provide a general interpretation of the results in the context of other evidence, and implications for future research. | 19 |
| **FUNDING** | | | |
| Funding | 27 | Describe sources of funding for the systematic review and other support (e.g., supply of data); role of funders for the systematic review. | 1 |

From: Moher D, Liberati A, Tetzlaff J, Altman DG, The PRISMA Group (2009). Preferred Reporting Items for Systematic Reviews and Meta-Analyses: The PRISMA Statement. PLoS Med 6(7): e1000097. doi:10.1371/journal.pmed1000097

| Item No | Recommendation | Reported on Page No |
|---|---|---|
| Reporting of background should include | | |
| 1 | Problem definition | 4 |
| 2 | Hypothesis statement | - |
| 3 | Description of study outcome(s) | 7 |
| 4 | Type of exposure or intervention used | 5 |
| 5 | Type of study designs used | 5-7 |
| 6 | Study population | 5-7 |
| Reporting of search strategy should include | | |
| 7 | Qualifications of searchers (eg, librarians and investigators) | Title page |
| 8 | Search strategy, including time period included in the synthesis and key words | 5, Appendix S2 |
| 9 | Effort to include all available studies, including contact with authors | 5,6,8 |
| 10 | Databases and registries searched | 5 |
| 11 | Search software used, name and version, including special features used (eg, explosion) | - |
| 12 | Use of hand searching (eg, reference lists of obtained articles) | 5 |
| 13 | List of citations located and those excluded, including justification | 10, Appendix S3, Table 1 |
| 14 | Method of addressing articles published in languages other than English | - |
| 15 | Method of handling abstracts and unpublished studies | - |
| 16 | Description of any contact with authors | 6,8 |
| Reporting of methods should include | | |
| 17 | Description of relevance or appropriateness of studies assembled for assessing the hypothesis to be tested | 10, Table 1, Table S2 |
| 18 | Rationale for the selection and coding of data (eg, sound clinical principles or convenience) | 6 |
| 19 | Documentation of how data were classified and coded (eg, multiple raters, blinding and interrater reliability) | 6 |
| 20 | Assessment of confounding (eg, comparability of cases and controls in studies where appropriate) | 6-8 |
| 21 | Assessment of study quality, including blinding of quality assessors, stratification or regression on possible predictors of study results | 7, 10, Table S3 and S4 |
| 22 | Assessment of heterogeneity | 11-13, Appendix S4 and S5 |
| 23 | Description of statistical methods (eg, complete description of fixed or random effects models, justification of whether the chosen models account for predictors of study results, dose- | 7-9 |

| | | |
|---|---|---|
| | response models, or cumulative meta-analysis) in sufficient detail to be replicated | |
| 24 | Provision of appropriate tables and graphics | Tables 1-2, S1-7, Figures 1-3, Appendix S4 |
| Reporting of results should include | | |
| 25 | Graphic summarizing individual study estimates and overall estimate | Figures 1-3, Appendix S4 |
| 26 | Table giving descriptive information for each study included | Tables 1-2 |
| 27 | Results of sensitivity testing (e.g, subgroup analysis) | 11-13, Appendix S5, Table S7 |
| 28 | Indication of statistical uncertainty of findings | 11-13, Figures 1-3, Appendix S4-5 |

From: Stroup DF, Berlin JA, Morton SC, et al, for the Meta-analysis Of Observational Studies in Epidemiology (MOOSE) Group. Meta-analysis of Observational Studies in Epidemiology. A Proposal for Reporting. JAMA. 2000;283(15):2008-2012. doi: 10.1001/jama.283.15.2008.

**SUPPLEMENTARY APPENDIX 2: Search string for the MEDLINE (OVID) database**

The search was conducted in December 2020.

1. Acute/ OR semi-acute/ OR urgent/ OR semi-urgent/ OR semi urgent/ OR emergen*/ OR semi-emergen*
2. (Laparoscop*/ OR minimally invasive/ OR keyhole/ OR surgery/ OR anaesthesia/ OR anesthesia) AND cholecystectomy/
3. 1 AND 2
4. Night/ OR Night-time/ OR Nighttime/ OR out-of-hour*/ OR after-hour*/ OR off-hour*/ OR hour*/ OR tim*/ OR week*
5. Day/ OR day-time
6. 4 OR 5
7. 3 AND 6

**TABLE S1: List of extracted data from included studies**

| Grouping | Data points |
|---|---|
| Study characteristics | First author, publication year, country, number of included centres or institutions, study period, study design |
| Definitions and criteria | Inclusion criteria, diagnostic criteria, exclusion criteria, out-of-hours and in-hours definitions |
| Surgical service characteristics | Number of hospital beds, hours of operation, acute surgical unit (yes/no), total number of operating theatres, operating theatres dedicated to emergency surgery (yes/no), number of available out-of-hours operating theatres, who performed the out-of-hours cholecystectomies |
| Pre-operative/patient demographic variables | Number of patients, age, sex, ASA classification (I-V), diagnosis, Tokyo severity grade, time from hospital admission to cholecystectomy |
| Planned operative approach | Open or laparoscopic |
| Intra-operative variables | Operative time, blood loss, conversion to open cholecystectomy, whether IOC was used, whether the critical view of safety was obtained |
| Post-operative complication rates | Any complication(s) according to the CD classification, biliary complications (bile leakage and/or BDI), wound infection, intra-abdominal abscesses, bleeding, sepsis, pneumonia, readmission, mortality |

ASA, American Society of Anesthesiologists; BDI, bile duct injury; CD, Clavien-Dindo; IOC, intra-operative cholangiography.

**SUPPLEMENTARY APPENDIX 3: PRISMA flow diagram outlining the selection process for included studies**

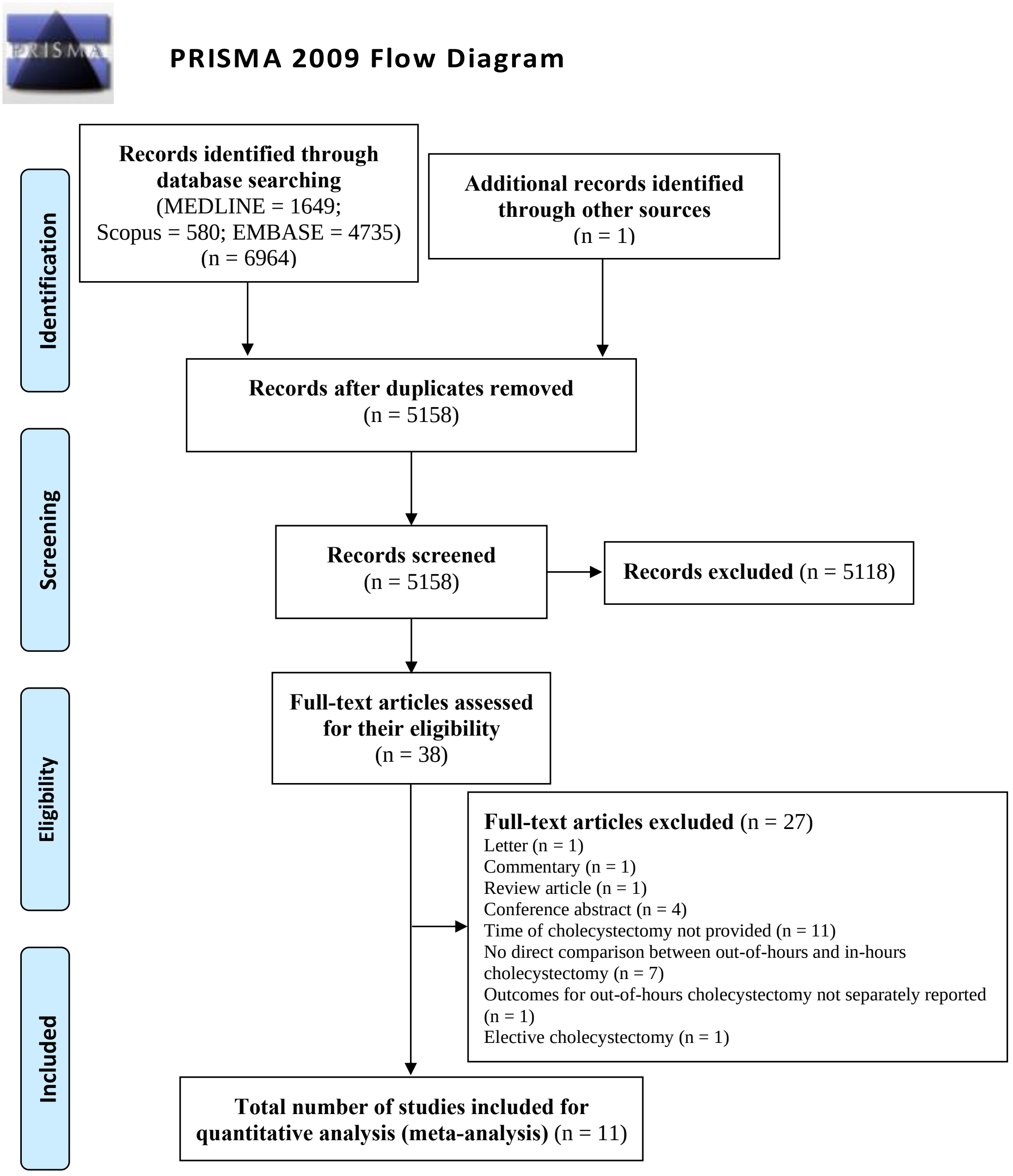

*From:* Moher D, Liberati A, Tetzlaff J, Altman DG, The PRISMA Group (2009). *P*referred *R*eporting *I*tems for *S*ystematic Reviews and *M*eta-*A*nalyses: The PRISMA Statement. PLoS Med 6(7): e1000097. doi:10.1371/journal.pmed1000097

**For more information, visit www.prisma-statement.org.**

#### TABLE S2: Selection criteria and out-of-hours definitions in included studies

| Author | Inclusion criteria | Exclusion criteria | Diagnostic criteria | Out-of-hours definition | In-hours definition |
|---|---|---|---|---|---|
| Geraedts [11] | Patients who underwent LC [a] | | N/A | Weekends, national holidays and daily (weekdays) between 5 PM and 8 AM. | Daily (weekdays) between 8 AM and 5 PM. |
| Gustafsson [12] | Patients undergoing non-elective cholecystectomy (for which time of day was recorded in >1% of procedures) for AC | Procedures for which either the time of day was not recorded or where only the duration of the procedure (in minutes) was recorded | | Procedures beginning between 19.00 and 07.00 hours (weekdays) or at any time during the weekend (from Friday 19.00 hours until Monday 07.00 hours) | Procedures beginning between 07.00 hours and 19.00 on weekdays. |
| Hoehn [27] | Patients undergoing urgent or emergent cholecystectomy; cholecystectomy procedures were identified based on the ICD-9 code (51.2x) | Patients with elective or trauma admissions | | Cholecystectomies performed on the weekend | Cholecystectomies performed on a weekday |
| Li [31] | Patients who underwent LC for AC during the same admission | Patients with concomitant gallstone pancreatitis and CBD stones | AC was defined by clinical criteria: acute onset right upper quadrant pain, with fever above 37.5$^{o}$C and/or elevated leukocyte count of $10x10^{9}$/L and imaging findings (i.e. gallstones and/or signs of inflammation on USS or CT scan) | LC operation start time later than 20.00 hours and earlier than 08.00 hours | LC operation start time later than 08.00 hours and earlier than 20.00 hours |
| Phatak [9] | Patients who underwent non-elective LC | | N/A | LC operations performed between 11 PM and 7 AM | LC operations performed between 7 AM and 11 PM |
| Rothman [29] | Adults (≥18 years) who underwent cholecystectomy by either open surgery or standard four-port laparoscopy | Patients who underwent open cholecystectomy in accordance with Danish cholecystectomy guidelines; Patients treated with single-incision cholecystectomy (SILS) or natural orifice transluminal endoscopic surgery (NOTES) technique; Patients with missing data on operation type (open, laparoscopic, SILS, NOTES), type of admission (emergency or elective) or other risk factors used for adjusting the outcome | AC was defined according to clinical and ultrasonic preoperative findings combined with perioperative findings of gallbladder wall edema; Chronic cholecystitis required a thickened wall with fibrosis and dense adhesions (judged by the operating surgeon); Previous pancreatitis was defined as an admission with the diagnosis of pancreatitis 3 months prior to surgery; Previous upper abdominal surgery was defined | Weekend days (Saturday, Sunday and public holidays occurring on weekdays) | Weekdays (Monday to Friday) |

| | | | | | |
|---|---|---|---|---|---|
| | | | as a scar in the area between the umbilicus and xiphoid process | | |
| Siada [6] | Patients who were admitted from the ED to the acute care surgery service with a diagnosis of AC and subsequently underwent LC | Patients who underwent elective cholecystectomy, incidental cholecystectomy, planned open cholecystectomy, had gallstone pancreatitis or diagnosed choledocholithiasis and those admitted to the medicine service | | Operation start times scheduled after 5 PM | Operations tart times scheduled until 5 PM |
| Tseng [13] | Patients with inpatient or emergency status who were booked for non-elective LC for any indication | Patients who underwent elective LC at the outpatient surgery centre; Patients whose cases were booked as open cholecystectomies | N/A | Operations starting between 7 PM and 6.59 AM | Operations starting between 7 AM and 6.59 PM |
| Wu [10] | Patients who underwent LC with a preoperative diagnosis of AC; Patients with suspected choledocholithiasis were included if they did not require an ERCP or if the ERCP was found to be negative for CBD stones | Patients with concomitant complicated gallstone disease (ERCP-confirmed choledocholithiasis, gallstone pancreatitis, ascending cholangitis) and previous insertion of a cholecystostomy tube; Patients with conditions (i.e. BDI, cystic duct stump leak, biloma, retained stone, SSI, pneumonia, UTI, myocardial infarction, cardiac arrhythmia, respiratory failure, DVT, PE, AKI, TIA/cerebrovascular accident) present on admission | | LC performed between 7 PM to 7 AM | LC performed between 7 AM to 7 PM |
| Yoshioka [32] | Patients diagnosed with AC who had been symptomatic for over 3 days and those with symptom onset within 72 hours, undergoing LC | Patients exhibiting poor general condition with regards to cardiopulmonary, liver and renal function, liver abscesses or biliary peritonitis; Patients receiving intensive antithrombotic therapy and those with concomitant CBD stones | AC diagnosed in accordance with the Revised Tokyo Guidelines [14] | LC operations starting at any time on a weekend day (Saturday or Sunday), between 9 PM and 9 AM on a weekday (Monday to Friday) or on a statutory holiday (in Japan) | LC operations starting between 9 AM and 9 PM on a weekday (Monday to Friday) |
| Zapf [28] | Inpatient stays for patients who underwent emergent or urgent cholecystectomy (performed on | | | Hospital admission on a weekend | Hospital admission on a weekday |

| | |
|---|---|
| | the same day as admission) for AC. Cholecystectomy procedures were identified according to ICD-9 codes |

AC, acute cholecystitis; AKI, acute kidney injury; BDI, bile duct injury; CBD, common bile duct; CT, computed tomography; DVT, deep vein thrombosis; ED, emergency department; ERCP, endoscopic retrograde cholangiopancreatography; ICD, International Classification of Diseases; LC, laparoscopic cholecystectomy; N/A, not applicable; PE, pulmonary embolism; SSI, surgical site infection; TIA, transient ischemic attack; USS, ultrasound scan; UTI, urinary tract infection.
Blank cells relate to data points that were not specified in the respective article.

[a] A subset of patients who underwent elective LC were also included; however, only urgent LC were analyzed.

**TABLE S3: Structure and characteristics of each surgical service**

| Author | No. of hospital beds | Hours of operation | Acute surgical unit (Yes/No) | No. of operating theatres (total) | Operating theatres dedicated to emergency surgery (Yes/No) | No. of available out-of-hours operating theatres | Who performed out-of-hours cholecystectomies |
|---|---|---|---|---|---|---|---|
| Geraedts [11] | | | | | | | |
| Gustafsson [12, a] | N/A | N/A | N/A | N/A | N/A | N/A | N/A |
| Hoehn [27, a] | N/A | N/A | N/A | N/A | N/A | N/A | N/A |
| Li [31] | | | | | | | Higher surgical trainee under supervision and/or specialist in general surgery |
| Phatak [9] | 328 | 24/7 | Yes | 8 (7AM-3PM); 4 (3PM-7PM); 2 (7PM-11PM); 1 (11PM-7AM) | No | 1 | Fellows (nighttime & weekend daytime); Junior & Senior (all day and nights of the week) |
| Rothman [29, a] | N/A | N/A | N/A | N/A | N/A | N/A | N/A |
| Siada [6] | 650 | 24/7 | Yes | | Yes (at least one) | | General surgery residents, acute surgical care and critical care fellows, advanced practice providers and attending surgeons |
| Tseng [13] | | | Yes | | Yes (at least one) | | General surgery residents, surgical critical care fellows and attending surgeons |
| Wu [10] | | | | | | | |

| | | | | | | | |
|---|---|---|---|---|---|---|---|
| Yoshioka [32] | | | | | | | Surgeons with >10 years of experience with LC surgery |
| Zapf [28, a] | N/A | N/A | N/A | N/A | N/A | N/A | N/A |

LC, laparoscopic cholecystectomy; N/A, not applicable.
Blank cells corresponding with data points that were not specified in their respective study.

[a] Data were obtained from large registries or population-based databases and therefore structure and characteristics of each surgical service pertaining to each individual centre or institution could not be obtained.

**TABLE S4: Methodological quality assessment of the included studies using the Newcastle-Ottawa Quality Assessment Scale**

| Authors | Quality assessment domains | | | | | | | | Total (/9★) |
|---|---|---|---|---|---|---|---|---|---|
| | Study group selection | | | | Comparability* | Ascertainment of outcome | | | |
| | Representativeness (★) | Selection of non-exposed cohort (★) | Ascertainment of exposure (★) | Demonstration that outcome was not present at study commencement (★) | Comparability of cohorts (★★) | Assessment of outcome (★) | Follow-up duration (★) | Adequacy of follow-up (★) | |
| Geraedts 2018 [11] | ★ | ★ | ★ | ★ | ★★ | ★ | ★ | ★ | 9★ |
| Gustafsson 2020 [12] | ★ | ★ | ★ | ★ | ★★ | ★ | ★ | | 8★ |
| Hoehn 2018 [27] | ★ | ★ | ★ | ★ | ★★ | ★ | ★ | | 8★ |
| Li 2009 [31] | ★ | ★ | ★ | ★ | ★★ | ★ | ★ | ★ | 9★ |
| Phatak 2014 [9] | ★ | ★ | ★ | ★ | ★★ | ★ | ★ | | 8★ |
| Rothman 2016 [29] | ★ | ★ | ★ | ★ | ★★ | ★ | ★ | | 8★ |
| Siada 2017 [6] | ★ | ★ | ★ | ★ | | ★ | ★ | | 6★ |
| Tseng 2019 [13] | ★ | ★ | ★ | ★ | | ★ | ★ | | 6★ |
| Wu 2014 [10] | ★ | ★ | ★ | ★ | ★★ | ★ | ★ | | 8★ |
| Yoshioka 2019 [32] | ★ | ★ | ★ | ★ | | ★ | ★ | ★ | 7★ |
| Zapf 2015 [28] | ★ | ★ | ★ | ★ | ★★ | ★ | | | 7★ |

* Two stars may be awarded depending on the number of factors being controlled based on the study design or analysis, to enable comparability between the two cohorts. One star is awarded if only a single factor is controlled and two stars are awarded if two or more factors are controlled.

**TABLE S5: Quality appraisal of included studies using the JBI Critical Appraisal Checklist for Cohort Studies**

| Author | Appraisal Question | | | | | | | | | | |
|---|---|---|---|---|---|---|---|---|---|---|---|
| | **1** | **2** | **3** | **4** | **5** [a] | **6** | **7** | **8** | **9** | **10** | **11** |
| Geraedts [11] | Y | Y | Y | Y | Y | Y | Y | Y | N/A | N/A | Y |
| Gustafsson [12] | Y | Y | Y | Y | Y | Y | Y | Y | N/A | N/A | Y |
| Hoehn [27] | Y | Y | Y | Y | Y | Y | U | Y | N/A | N/A | Y |
| Li [31] | Y | Y | Y | Y | Y | Y | U | N/A | N/A | N/A | Y |
| Phatak [9] | Y | Y | Y | Y | Y | Y | U | Y | N/A | N/A | Y |
| Rothman [29] | Y | Y | Y | Y | Y | Y | Y | Y | N/A | N/A | Y |
| Siada [6] | Y | Y | Y | N | N | Y | Y | Y | N/A | N/A | Y |
| Tseng [13] | Y | Y | Y | N | N | Y | Y | Y | N/A | N/A | Y |
| Wu [10] | Y | Y | Y | Y | Y | Y | Y | Y | N/A | N/A | Y |
| Yoshioka [32] | Y | Y | Y | Y | N | Y | Y | U | N/A | N/A | Y |
| Zapf [28] | Y | Y | Y | Y | Y | Y | Y | U | N/A | N/A | Y |

JBI, Joanna Briggs Institute; N, No; N/A, Not Applicable; U, Unclear; Y, Yes.

[a] Covariates included within the adjusted multivariate analysis model for each study are shown in **Table S6.**

**Appraisal questions:**

1. Were the two groups similar and recruited from the same population?
2. Were the exposures measured similarly to assign people to both exposed and unexposed groups?
3. Was the exposure measured in a valid and reliable way?
4. Were confounding factors identified?
5. Were strategies to deal with confounding factors stated?
6. Were the groups/participants free of the outcome at the start of the study (or at the moment of exposure)?
7. Were the outcomes measured in a valid and reliable way?
8. Was the follow up time reported and sufficient to be long enough for outcomes to occur?
9. Was follow up complete, and if not, were the reasons to loss to follow up described and explored?

10. Were strategies to address incomplete follow up utilized?
11. Was appropriate statistical analysis used?

#### TABLE S6: Outcome definitions provided by each study

| **Author** | **Outcome and definition** | | |
|---|---|---|---|
| | **Post-operative complication** | **Intra-operative complications** | **Readmission** |
| Geraedts [11] | Any morbidity during the same hospitalization or within 90 days of surgery | | |
| Gustafsson [12] | Any complication (yes/no) within 30 days | | |
| Hoehn [27] | | | |
| Li [31] | | | |
| Phatak [9] | Bile leak/biloma, CBD injury, retained bile duct stone, organ space abscess, superficial SSI, bleeding requiring reoperation, and pneumonia | | |
| Rothman [29] | | | |
| Siada [6] | Any complication within 30 days of the operation and includes: bile leak, biloma, retained CBD stone, cystic duct stump leak, intra-abdominal abscess, wound infection, cardiac complications (arrhythmias, arrest, MI), respiratory complications (respiratory failure, pneumonia, PE), DVT, hemorrhage, pancreatitis, AKI, and infrequent miscellaneous complications (upper GI bleeding, colitis, urosepsis) | Any of the following: vascular injury, hollow viscus injury and biliary injury | |
| Tseng [13] | | | Any reason for return to the hospital within 60 days of discharge other than for scheduled outpatient appointments |
| Wu [10] | Any of the following occurring within 30 days of hospitalisation: BDI necessitating endoscopy or surgical intervention, cystic duct stump leak, biloma requiring external drainage, retained bile duct stone, SSI (superficial/deep), pneumonia, UTI, MI, cardiac arrhythmia, respiratory failure requiring intubation, DVT or PE, AKI (defined as rise in creatinine by >0.3 mg/dL or 50% in 48 hours or urine output <0.5 mL/kg/hour for 6 hours) and TIA/cerebrovascular accident | | |
| Yoshioka [32] | Any complication rated ≥ grade 1 using the Clavien-Dindo classification system | | |
| Zapf [28] | The acquisition of an ICD-9 Clinical Modification code corresponding to the specific complication during the course of the stay that was not present upon admission to the hospital (wound complicaitons, blood transfusion, sepsis, pneumonia, UTI) | | |

AKI, acute kidney injury; BDI, bile duct injury; CBD, common bile duct; DVT, deep vein thrombosis; GI, gastrointestinal; ICD, International Classification of Diseases; MI, myocardial infarction; PE, pulmonary embolism; SSI, surgical site infection; TIA, transient ischemic attack; UTI, urinary tract infection;

Blank cells relate to definitions that were not provided.

**SUPPLEMENTARY APPENDIX 4: Meta-analyses and forest plots of outcomes of interest other than biliary complications, overall post-operative complications and conversion to open cholecystectomy following out-of-hours and in-hours urgent cholecystectomy**

Time from admission to cholecystectomy

Time from hospital admission to urgent cholecystectomy was reported in four studies [10,13,27,32]. Similar times were seen in out-of-hours and in-hours urgent cholecystectomy (MD -0.16 days, 95% CI: -0.91 to 0.60 days; p=0.69). There was considerable heterogeneity in the analyzed studies ($I^2$ = 99%).

| Study | Out-of-hours Total | Out-of-hours Mean | Out-of-hours SD | In-hours Total | In-hours Mean | In-hours SD | MD | 95%-CI | Weight |
|---|---|---|---|---|---|---|---|---|---|
| Hoehn 2018 | 22301 | 1.70 | 1.4826 | 111779 | 1.00 | 1.4827 | 0.70 | [ 0.68; 0.72] | 25.6% |
| Tseng 2019 | 576 | 1.00 | 1.4864 | 4628 | 1.35 | 0.7415 | -0.35 | [-0.47; -0.23] | 25.4% |
| Wu 2014 | 232 | 1.61 | 1.5666 | 908 | 2.07 | 1.4850 | -0.46 | [-0.69; -0.24] | 25.1% |
| Yoshioka 2019 | 101 | 2.04 | 1.6700 | 270 | 2.58 | 2.0000 | -0.54 | [-0.94; -0.14] | 23.9% |
| **Random effects model** | **23210** | | | **117585** | | | **-0.16** | **[-0.91; 0.60]** | **100.0%** |

Heterogeneity: $I^2$ = 99%
Test for overall effect: $z$ = -0.40 ($p$ = 0.69)

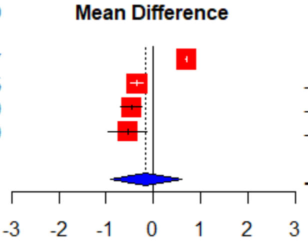

**Intra-operative**

Operative duration

Duration of urgent cholecystectomy were reported in six studies [6,9-11,13,32]. The operative duration were similar between out-of-hours *vs.* in-hours urgent cholecystectomy (MD +1.97 minutes, 95% CI: -8.37 to 12.66 minutes; p=0.72). There was considerable heterogeneity among the analyzed studies ($I^2$ = 97%).

| Study | Out-of-hours Total | Out-of-hours Mean | Out-of-hours SD | In-hours Total | In-hours Mean | In-hours SD | MD | 95%-CI | Weight |
|---|---|---|---|---|---|---|---|---|---|
| Geraedts 2018 | 145 | 64.50 | 36.2000 | 67 | 73.30 | 28.7000 | -8.80 | [-17.85; 0.25] | 15.7% |
| Phatak 2014 | 108 | 80.80 | 31.6000 | 248 | 81.90 | 33.2000 | -1.10 | [ -8.35; 6.15] | 16.4% |
| Siada 2017 | 219 | 102.00 | 43.0000 | 647 | 102.00 | 51.0000 | 0.00 | [ -6.92; 6.92] | 16.5% |
| Tseng 2019 | 576 | 93.80 | 37.9000 | 4628 | 95.60 | 34.1000 | -1.80 | [ -5.05; 1.45] | 17.4% |
| Wu 2014 | 232 | 110.50 | 12.2000 | 908 | 92.40 | 4.5000 | 18.10 | [ 16.50; 19.70] | 17.7% |
| Yoshioka 2019 | 101 | 95.00 | 33.0000 | 270 | 91.00 | 34.0000 | 4.00 | [ -3.61; 11.61] | 16.3% |
| **Random effects model** | **1381** | | | **6768** | | | **1.97** | **[ -8.73; 12.66]** | **100.0%** |

Heterogeneity: $I^2$ = 97%
Test for overall effect: $z$ = 0.36 ($p$ = 0.72)

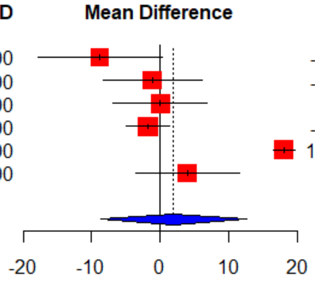

#### Blood loss

Intra-operative blood loss during urgent cholecystectomy were reported in three studies [11,13,32]. Significantly higher quantities of blood loss were observed during out-of-hours *vs.* in-hours urgent cholecystectomy (MD +1.49 mL, 95% CI: 0.21 to 2.78 mL; p=0.02). No heterogeneity was detected ($I^2$ = 0%).

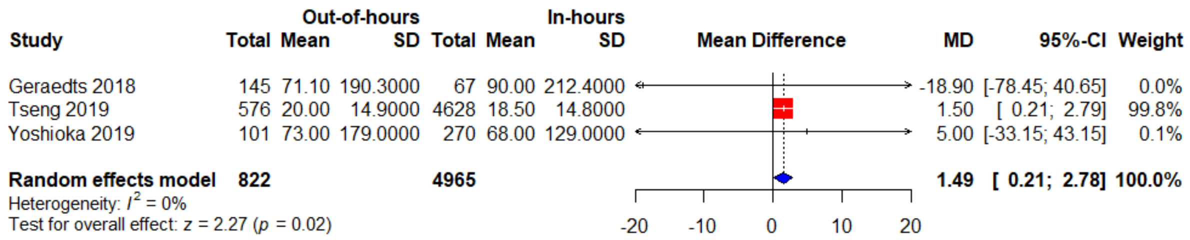

#### Intra-operative cholangiography

Two studies reported on the use of IOC during urgent cholecystectomy [6,9]. Significantly fewer IOCs were used during out-of-hours *vs.* in-hours urgent cholecystectomy (OR 0.68, 95% CI: 0.49 to 0.93; p=0.02). No heterogeneity was detected ($I^2$ = 0%).

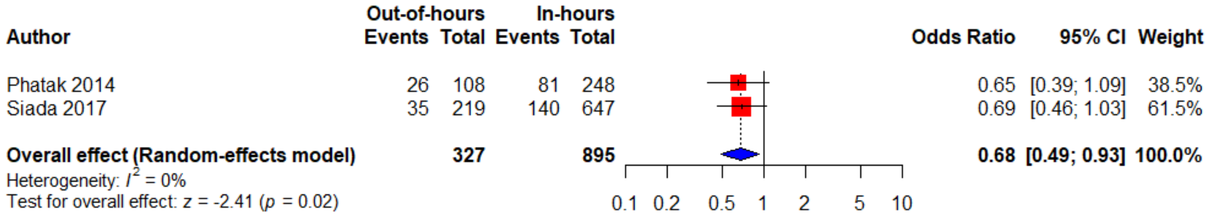

### Post-operative

#### Length of stay

Post-operative LOS after out-of-hours *vs.* in-hours urgent cholecystectomy were reported in six studies [9,10,13,27,28,32]. Post-operative LOS did not differ between out-of-hours *vs.* in-hours urgent cholecystectomy on univariate (MD -0.05 days, 95% CI: -0.55 to 0.45 days; p=0.30). and multivariate (MD . There was considerable heterogeneity between the analyzed studies

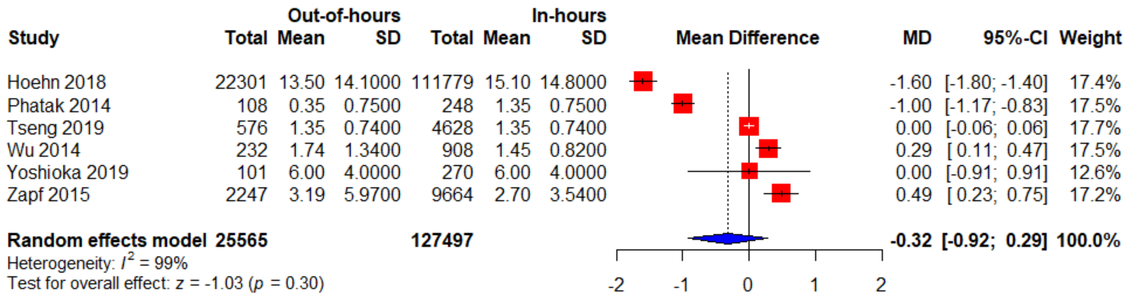

($I^2$ = 97%).

#### Wound infection

Wound infection following out-of-hours *vs.* in-hours urgent cholecystectomy was reported in three studies [9,28,32]. Wound infection rates were similar between out-of-hours *vs.* in-hours urgent cholecystectomy on both univariate (OR 1.41, 95% CI: 0.81 to 2.44; p=0.22) and multivariate (OR 1.39, 95% CI: 0.61 to 3.20; p>0.05) meta-analysis. No heterogeneity was detected ($I^2$ = 0%).

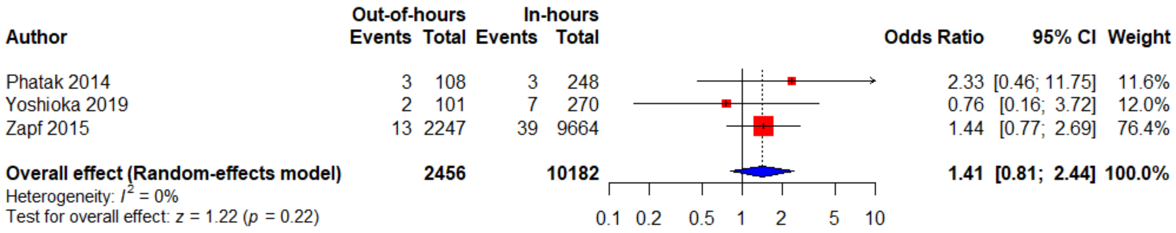

#### Intra-abdominal abscess

Two studies reported on the presence of an intra-abdominal abscess following urgent cholecystectomy [9,32]. Rates of intra-abdominal abscesses did not differ between out-of-hours *vs.* in-hours urgent cholecystectomy (OR 1.3, 95% CI: 0.22 to 7.57; p=0.77). No heterogeneity was detected ($I^2$ = 0%).

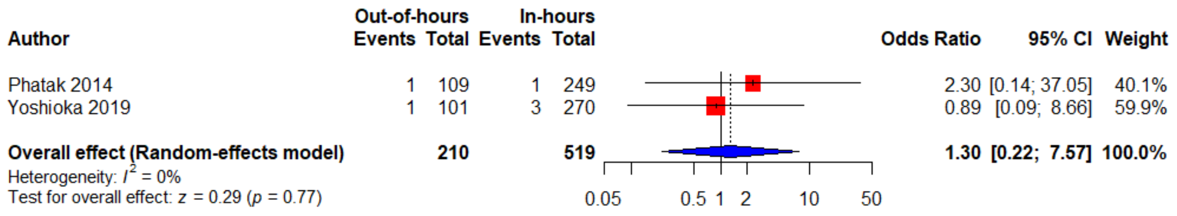

#### Bleeding

Post-operative bleeding following out-of-hours *vs.* in-hours urgent cholecystectomy was noted in two studies [9,32]. Rates of post-operative bleeding did not differ between out-of-hours *vs.* in-hours urgent cholecystectomy (OR 2.48, 95% CI; 0.45 to 13.7; p=0.30). No heterogeneity was detected ($I^2$ = 0%).

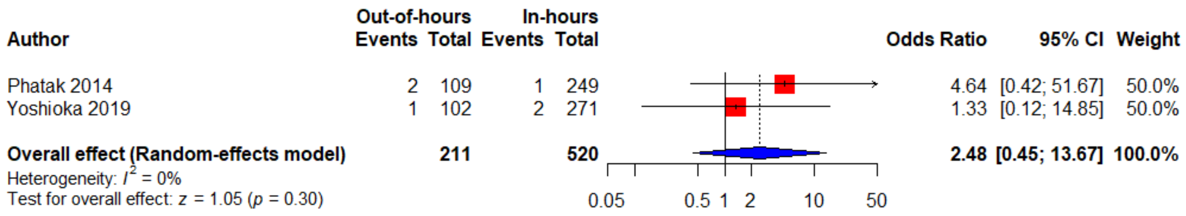

#### Sepsis

Rates of sepsis following out-of-hours *vs.* in-hours urgent cholecystectomy were reported in two studies [28,32]. Significantly higher rates of post-operative sepsis following out-of-hours *vs.* in-hours urgent cholecystectomy were observed on univariate (OR 1.58, 95% CI: 1.04 to 2.41; p=0.03) but not on multivariate (OR 0.81, 95% CI: 0.41 to 1.58; p>0.05) meta-analysis. No heterogeneity was detected ($I^2$ = 0%).

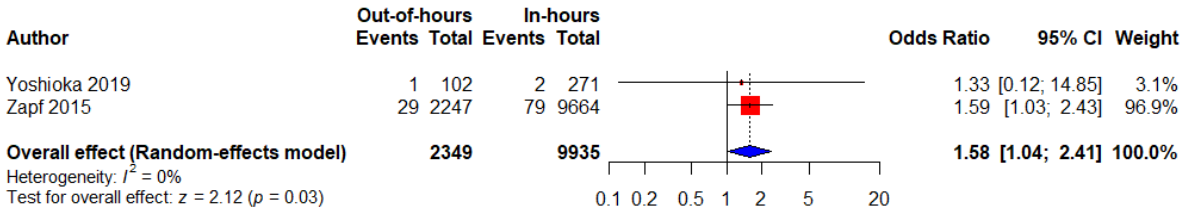

#### Pneumonia

Two studies reported pneumonia following out-of-hours *vs.* in-hours urgent cholecystectomy [28,32]. Significantly higher rates of pneumonia following out-of-hours *vs.* in-hours urgent cholecystectomy were observed on univariate (OR 1.55, 95% CI: 1.06 to 2.26; p=0.02) but not on multivariate (OR 1.10, 95% CI: 0.63 to 1.94; p>0.05) meta-analysis. No heterogeneity was detected ($I^2$ = 0%).

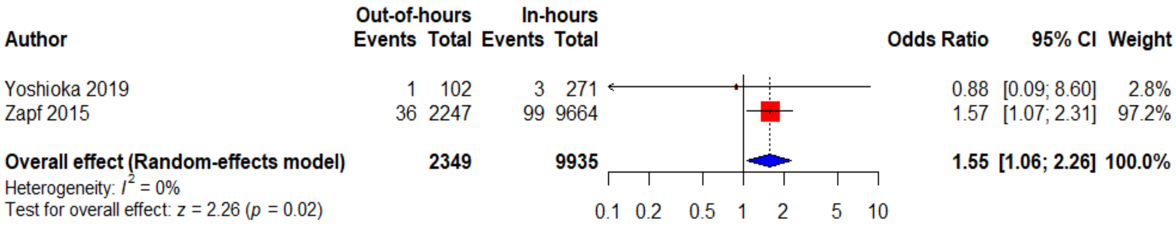

#### Readmission

Three studies reported on their readmission rates following out-of-hours *vs.* in-hours urgent cholecystectomy [13,27,29]. Readmission rates did not differ between out-of-hours *vs.* in-hours urgent cholecystectomy (OR 1.16, 95% CI: 0.96 to 1.40; p=0.73). There was considerable heterogeneity between the analyzed studies ($I^2$ = 88%).

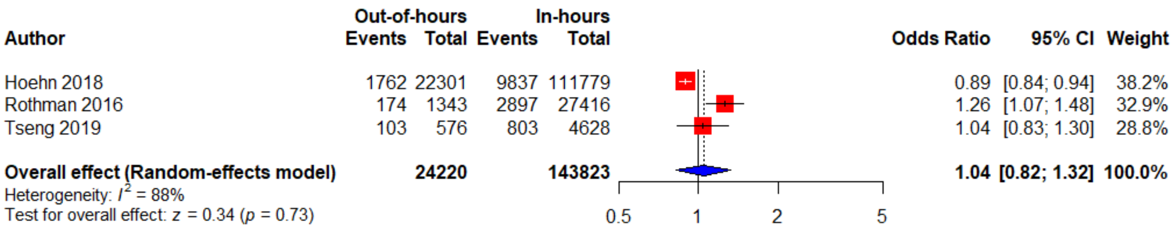

#### Mortality

Post-operative mortality rates were reported in seven studies [6,9,10,13,27,28,32]. Mortality rates following out-of-hours *vs.* in-hours urgent cholecystectomy did not differ (OR 1.38, 95% CI: 0.90 to 2.11; p=0.14). Heterogeneity was considered not important ($I^2$ = 33%).

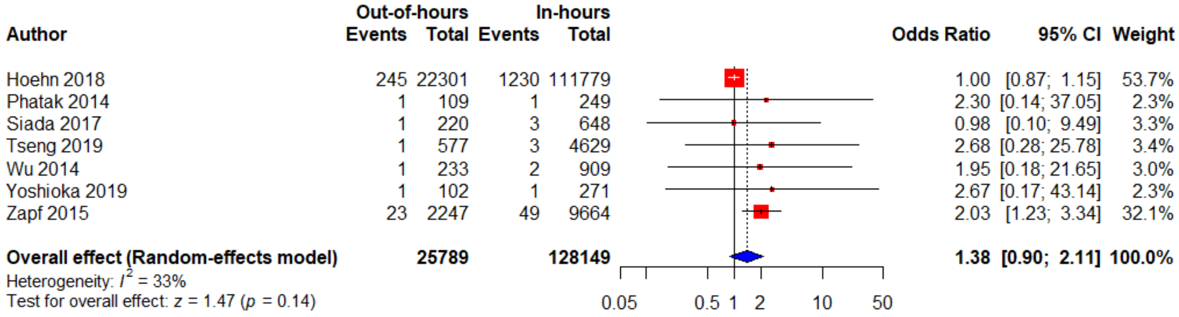

Total length of stay

Total hospital LOS were reported in two studies [9,10]. Total LOS was similar following out-of-hours and in-hours urgent cholecystectomy (MD -0.56 days, 95% CI: -1.41 to 0.28 days; p=0.19). There was considerable heterogeneity between the analyzed studies ($I^2$ = 93%).

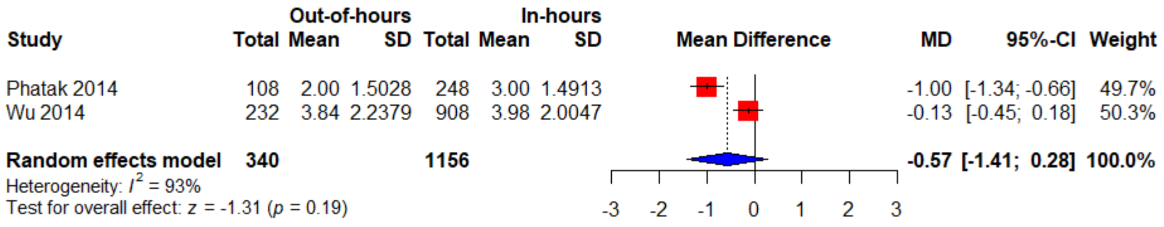

**SUPPLEMENTARY APPENDIX 5: Subgroup analyses of evenings and night-time *vs.* daytime, weekend *vs.* weekday and laparoscopic-only urgent cholecystectomy**

**Evenings and night-time *vs.* daytime urgent cholecystectomy**

| Outcome | No. of studies | References | OR (MD) | 95% CI | p-value* | $I^2$, % |
|---|---|---|---|---|---|---|
| Bile leakage | 1 | 9 | 1.15 | 0.019 – 22.30 | 1 | |
| Bile duct injury | 2 | 6, 9 | 1.06 | 0.20 – 5.52 | 0.95 | 0% |
| Post-operative complications | 3 | 6, 9, 10 | 0.93 | 0.58 – 1.52 | 0.79 | 29% |
| Conversion to open cholecystectomy | 4 | 6, 9, 10, 13 | 0.91 | 0.45 – 1.83 | 0.78 | 79% |
| Time from admission to cholecystectomy | 2 | 10, 13 | (-0.38) | -0.48 – -0.27 | **<0.0001** | 0% |
| Operative duration | 4 | 6, 9, 10, 13 | (3.97) | -9.20 - 17.13 | 0.55 | 98% |
| Intra-operative blood loss | 1 | 13 | (1.50) | 0.21 – 2.79 | **0.023** | |
| Post-operative LOS | 3 | 9, 10, 13 | (-0.24) | -0.88 – 0.41 | 0.47 | 99% |
| Wound infection | 1 | 9 | 2.33 | 0.46 – 11.75 | 0.30 | |
| Readmission | 1 | 13 | 1.04 | 0.83 – 1.30 | 0.75 | |

CI, confidence interval; LOS, length of stay; MD, mean difference; OR, odds ratio; p-value, probability value.
Empty cells relate to outcomes in which pooling via meta-analysis could not be performed.
*Statistically significant p-values (p<0.05) are bolded.

The rates of overall biliary complication, IOC, and total LOS did not differ on subgroup analysis as these outcomes were only reported in studies of night-time *vs.* daytime urgent cholecystectomy. Rates of post-operative pneumonia and sepsis were not reported in any studies of night-time *vs.* daytime urgent cholecystectomy and hence, subgroup analysis of these outcomes could not be performed. No instances of post-operative intra-abdominal abscesses, bleeding and mortality were observed in studies of night-time *vs.* daytime urgent cholecystectomy only and therefore, subgroup analysis of these outcomes could not be performed.

**Weekend *vs.* weekday urgent cholecystectomy**

| Outcome | No. of studies | References | OR (MD) | 95% CI | p-value * | $I^2$, % |
|---|---|---|---|---|---|---|
| Conversion to open cholecystectomy | 1 | 31 | 2.54 | 2.18 – 2.95 | **<0.0001** | |
| Time from admission to cholecystectomy | 1 | 27 | 0.70 | 0.68 – 0.72 | **<0.0001** | |
| Post-operative LOS | 2 | 27, 28 | (-0.56) | -2.60 – 1.49 | 0.59 | 99% |
| Wound infection | 1 | 28 | 1.43 | 0.77 – 2.69 | 0.26 | |
| Sepsis | 1 | 28 | 1.58 | 1.03 – 2.43 | **0.0346** | |
| Pneumonia | 1 | 28 | 1.57 | 1.07 – 2.31 | **0.0208** | |
| Readmission | 2 | 27, 29 | 1.05 | 0.75 – 1.48 | 0.78 | 94% |
| Mortality | 2 | 27, 28 | 1.37 | 0.68 – 2.72 | 0.38 | 86% |

CI, confidence interval; LOS, length of stay; MD, mean difference; OR, odds ratio; p-value, probability value.
Empty cells relate to outcomes in which pooling via meta-analysis could not be performed.
*Statistically significant p-values (p<0.05) are bolded.

Subgroup analysis of rates of overall biliary complications, bile leakage, BDI, post-operative complications, IOC, post-operative bleeding, intra-abdominal abscesses, operative duration, intra-operative blood loss, and total LOS could not be performed as they were not reported in any studies of weekend *vs.* weekday urgent cholecystectomy.

**Laparoscopic-only urgent cholecystectomy**

| Outcome | No. of studies | References | Pooled OR (MD) | 95% CI | p-value | $I^2$, % |
|---|---|---|---|---|---|---|
| Bile duct injury | 2 | 6, 9 | 1.06 | 0.20 – 5.52 | 0.95 | 0 |
| Post-operative complications | 4 | 6, 9, 10, 32 | 0.99 | 0.66 – 1.47 | 0.96 | 14 |
| Conversion to open cholecystectomy | 6 | 6, 9-11, 13, 32 | 1.09 | 0.62 – 1.90 | 0.77 | 69% |
| Post-operative LOS | 5 | 9, 10, 13, 27, 32 | (-0.49) | -1.17 – 0.20 | 0.17 | 99% |
| Wound infection | 2 | 9, 32 | 1.32 | 0.42 – 4.09 | 0.63 | 0% |
| Readmission | 2 | 13, 27 | 0.92 | 0.81 – 1.05 | 0.21 | 41% |
| Mortality | 6 | 6, 9, 10, 13, 27, 32 | 1.01 | 0.88 – 1.16 | 0.90 | 0% |

CI, confidence interval; LOS, length of stay; MD, mean difference; OR, odds ratio; p-value, probability value.

Rates of overall biliary complications, bile leakage, IOC, post-operative intra-abdominal abscesses, bleeding, time from admission to cholecystectomy, operative duration, intra-operative blood loss, and total LOS did not differ on subgroup analysis as they were only reported in studies of urgent cholecystectomy planned laparoscopically. Subgroup analysis of post-operative sepsis and pneumonia could not be performed as there were no reported instances of these outcomes.

**TABLE S7: Covariates included within the multivariate analysis for each study**

| Author | Adjusted covariates |
|---|---|
| Geraedts [11] | Out-of-hours, timing of surgery (emergency *vs.* elective), surgeon experience (senior *vs.* junior attending), conversion to laparotomy |
| Gustafsson [12] | Age (ordinal scale; <50, 50-75, >75), sex, ASA classification (dichotomised; 1-2 *vs.* 3-5), interval between admission and surgery (ordinal scale; 0, 1, 2, 3, 4-7, >8 days), hospital-specific procedures (proportion of procedures with a time of day registered per centre (<0.1, 0.1-0.3, 0.3-0.5, 0.5-0.7, >0.7) and proportion of procedures performed out-of-hours per centre (<0.1, 0.1-0.2, 0.2-0.3, 0.3-0.4, >0.4)) |
| Hoehn [27] | Age, sex, race, safety-net stratification (safety-net burden: proportion of all patient charges that were Medicaid or uninsured), severity of illness and procedure day (weekday or weekend) |
| Li [31] | Sex, time from admission to operation (>48h), duration of symptoms before admission, total bilirubin level, operation start time later than 20.00 hours and before 08.00 hours |
| Phatak [9] | Age, case duration, nighttime surgery |
| Rothman [29] | Age (<60 years), sex, ASA classification (<2), diagnosis (acute/chronic cholecystitis), previous upper abdominal surgery, surgical experience |
| Siada [6] | |
| Tseng [13] | |
| Wu [10] | Age, ASA classification, duration of acute symptoms before presentation, time from presentation to operation, operation time (night *vs.* day), Tokyo severity grade for acute cholecystitis [14] and gangrenous pathology |
| Yoshioka [32] | |
| Zapf [28] | Age, sex, race, insurance status, type of surgery (emergent *vs.* urgent), laparoscopic surgery (yes/no), pediatric (yes/no), number of chronic conditions, hospital size, profit status, location (urban *vs.* rural) |

ASA, American Society of Anesthesiologists; vs, versus.

#### SUPPLEMENTARY APPENDIX 6: Funnel plots for the outcomes of interest

*Overall biliary complications*

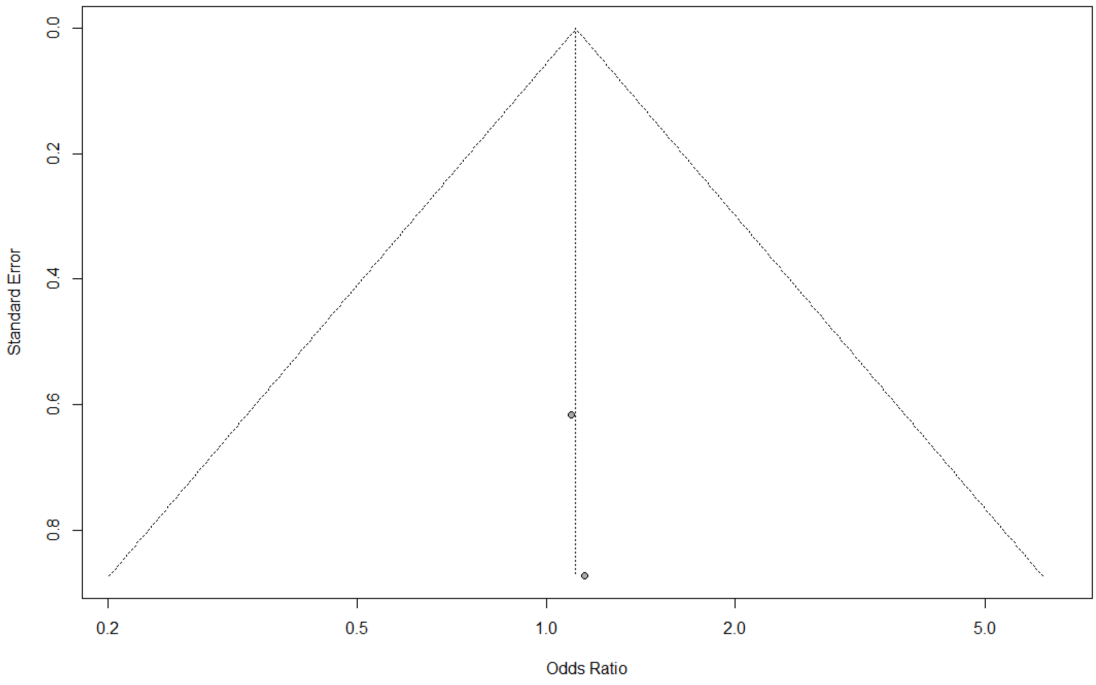

*Overall post-operative complications*

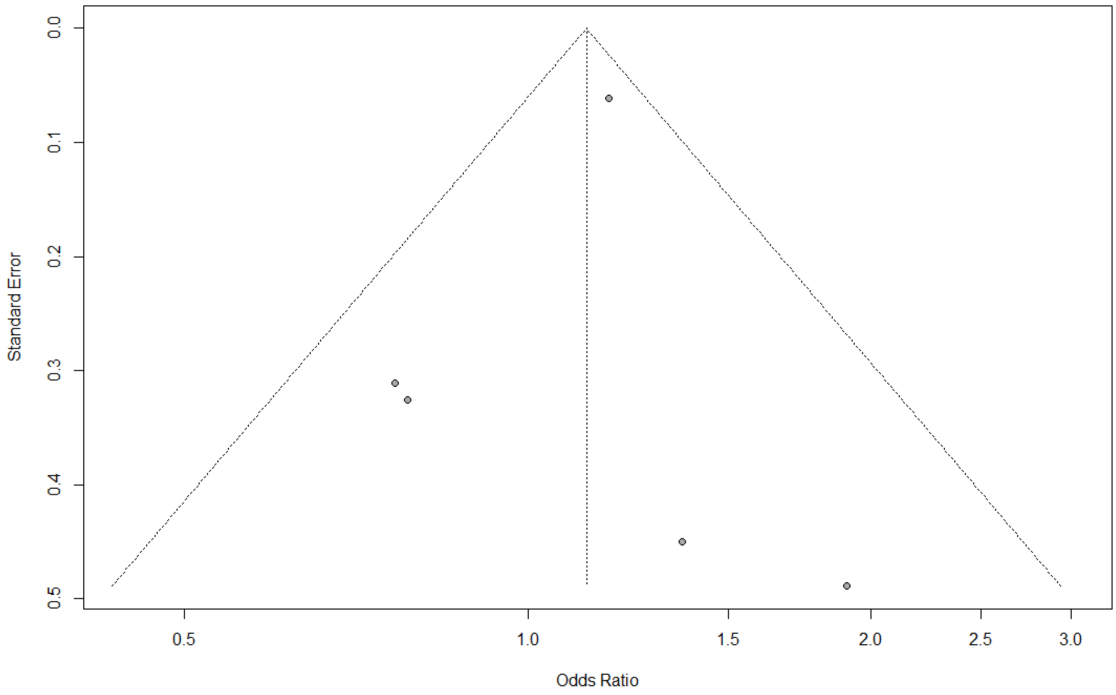

*Conversion to open cholecystectomy*

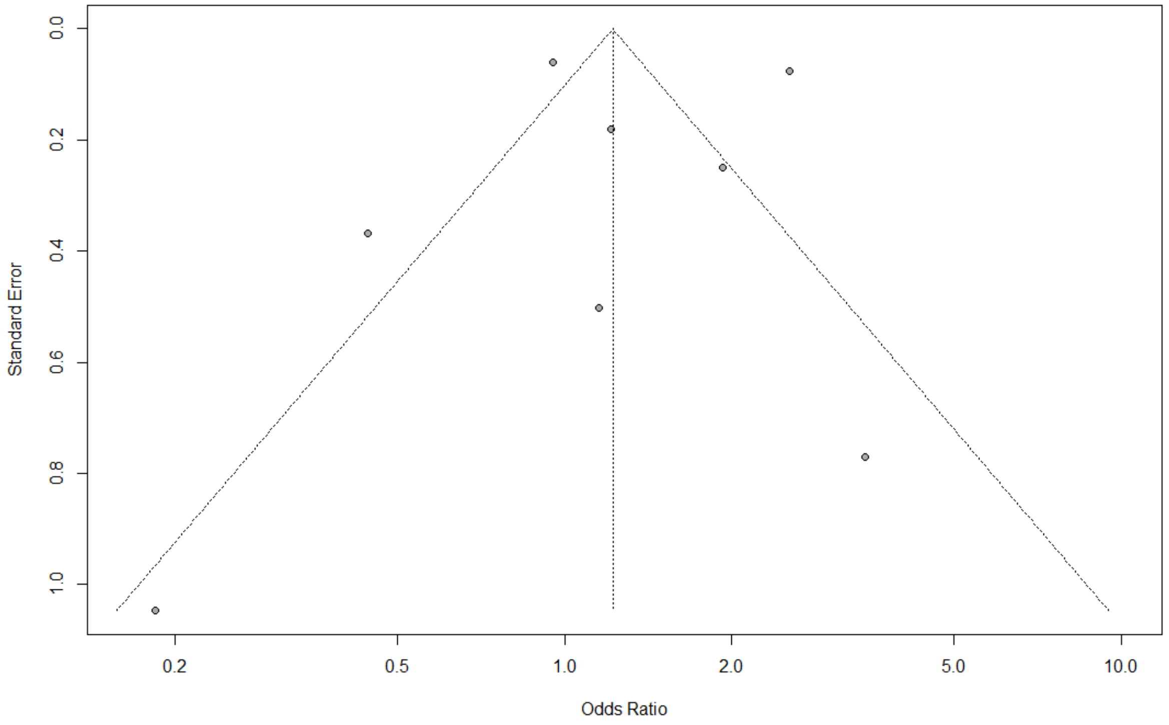

*Time to cholecystectomy*

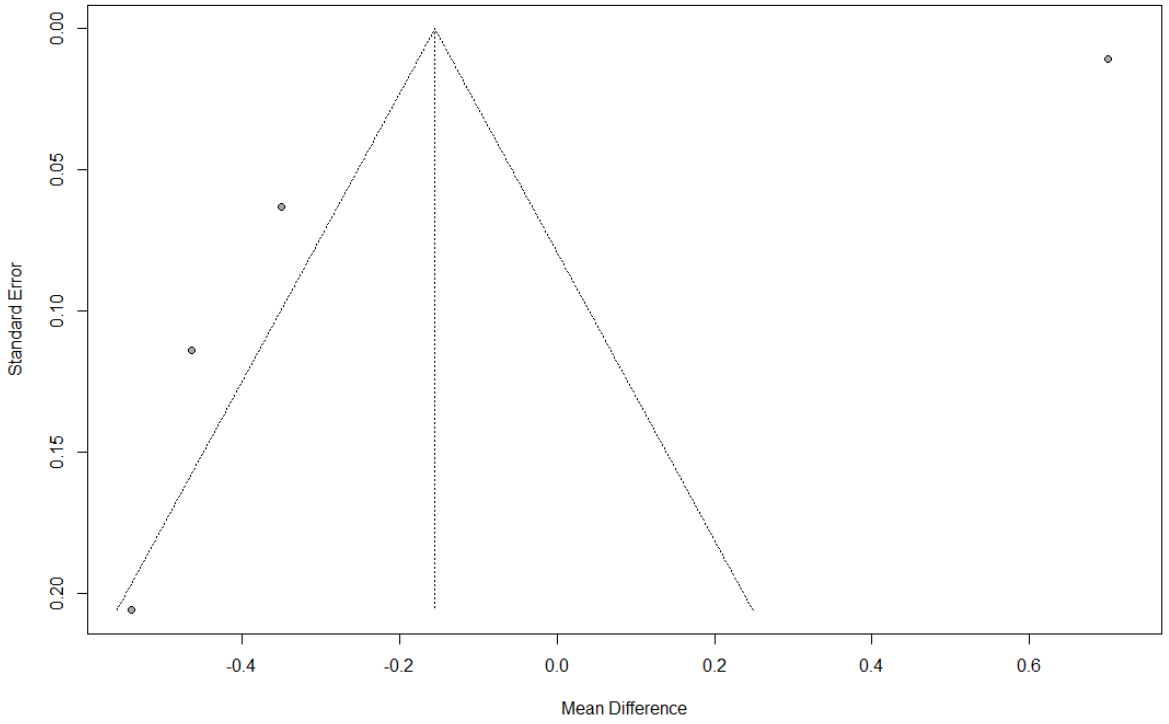

#### Intra-operative

*Operative duration*

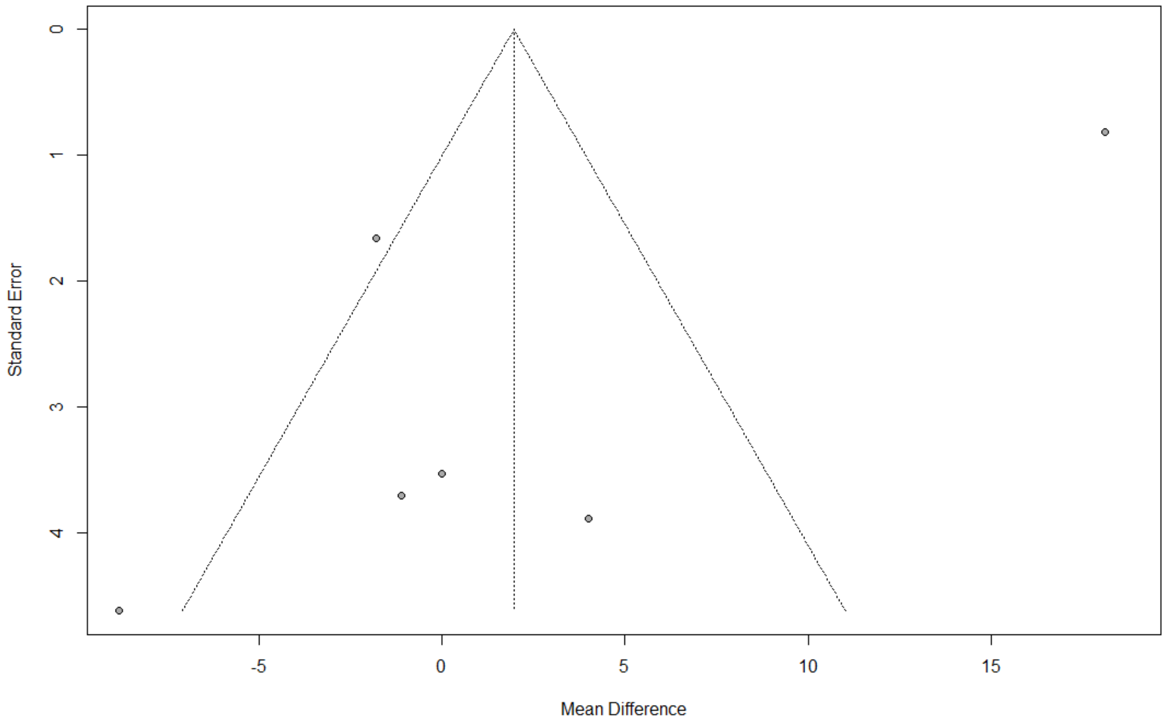

*Intra-operative blood loss*

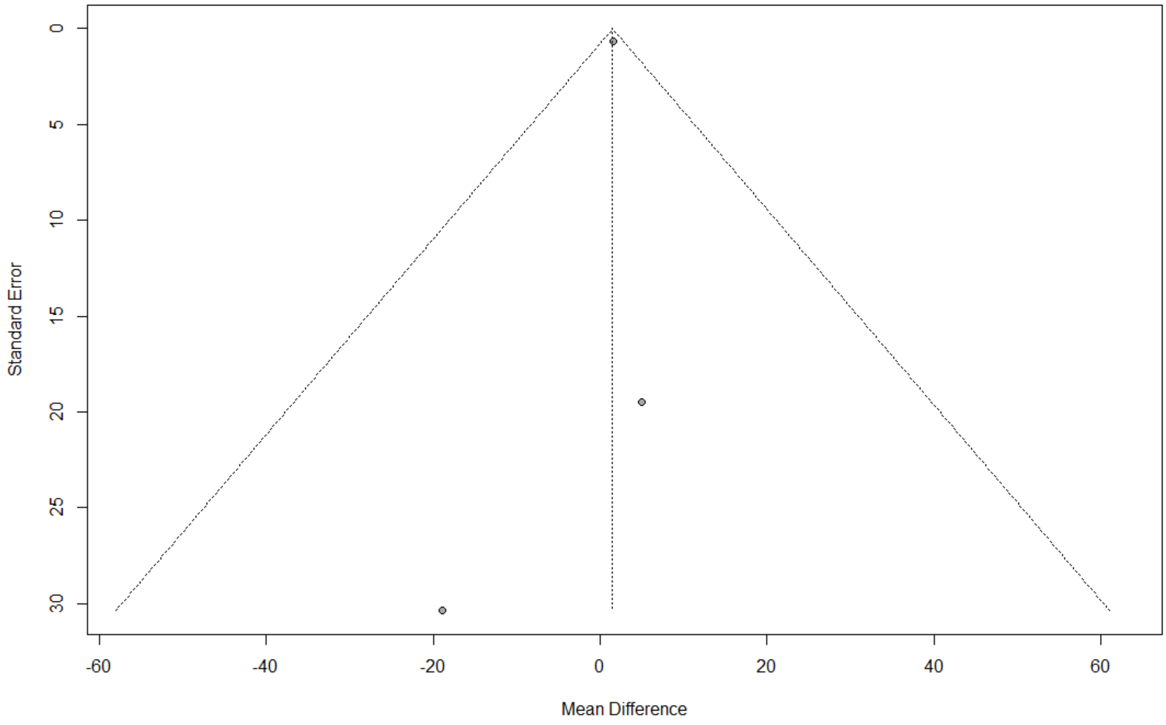

*Intra-operative cholangiography*

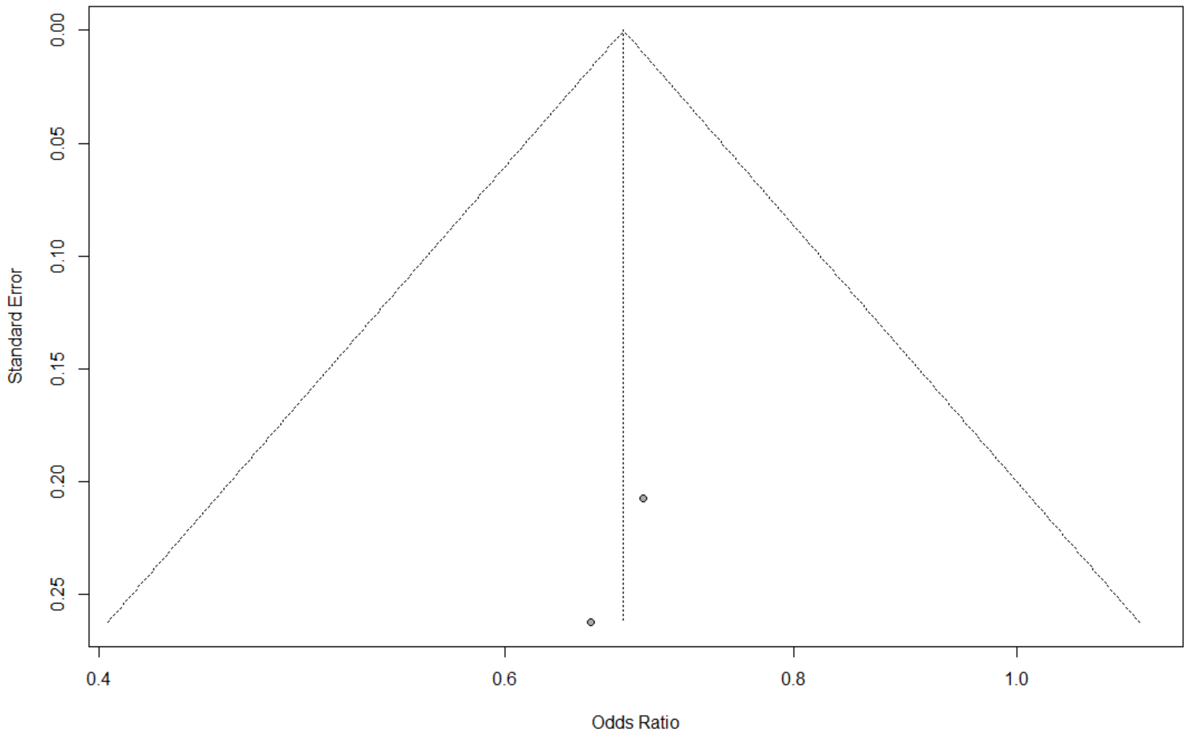

#### Post-operative

*Length of stay*

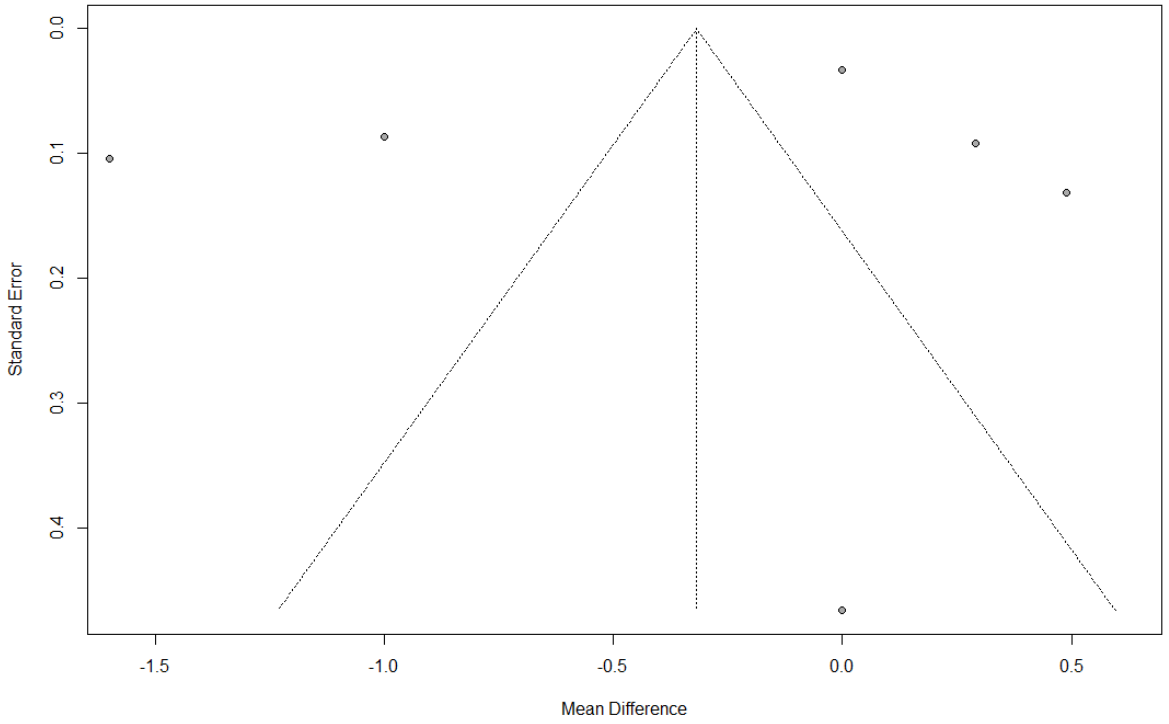

*Wound infection*

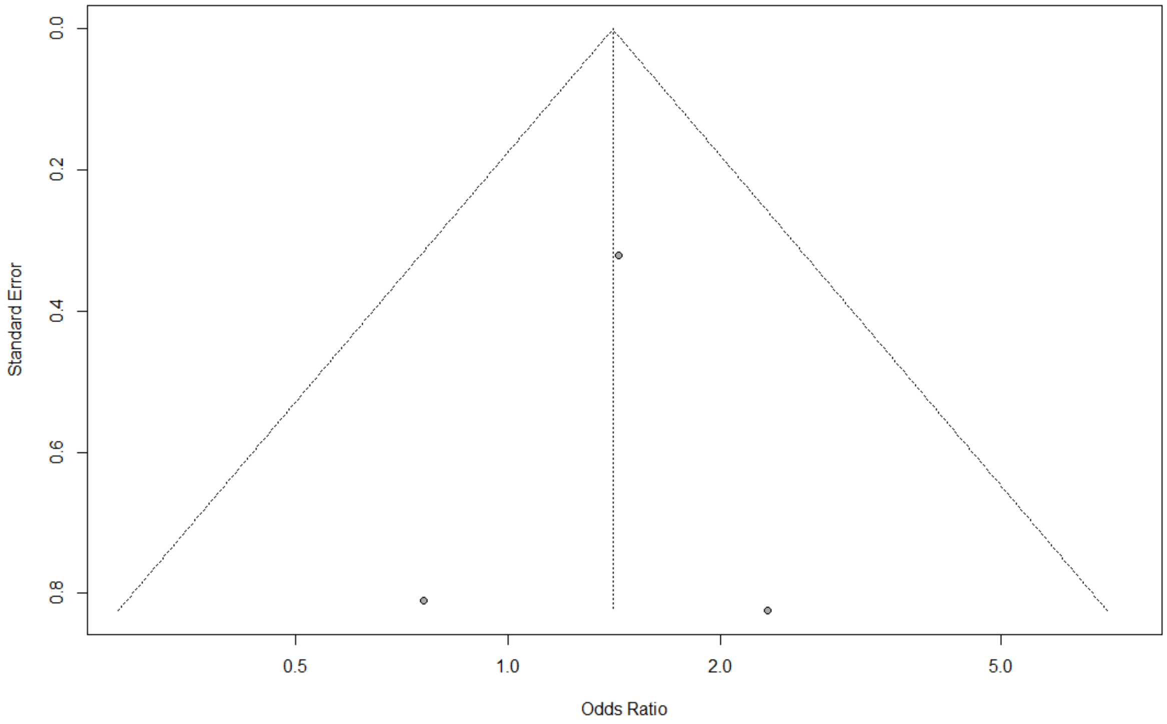

*Intra-abdominal abscess*

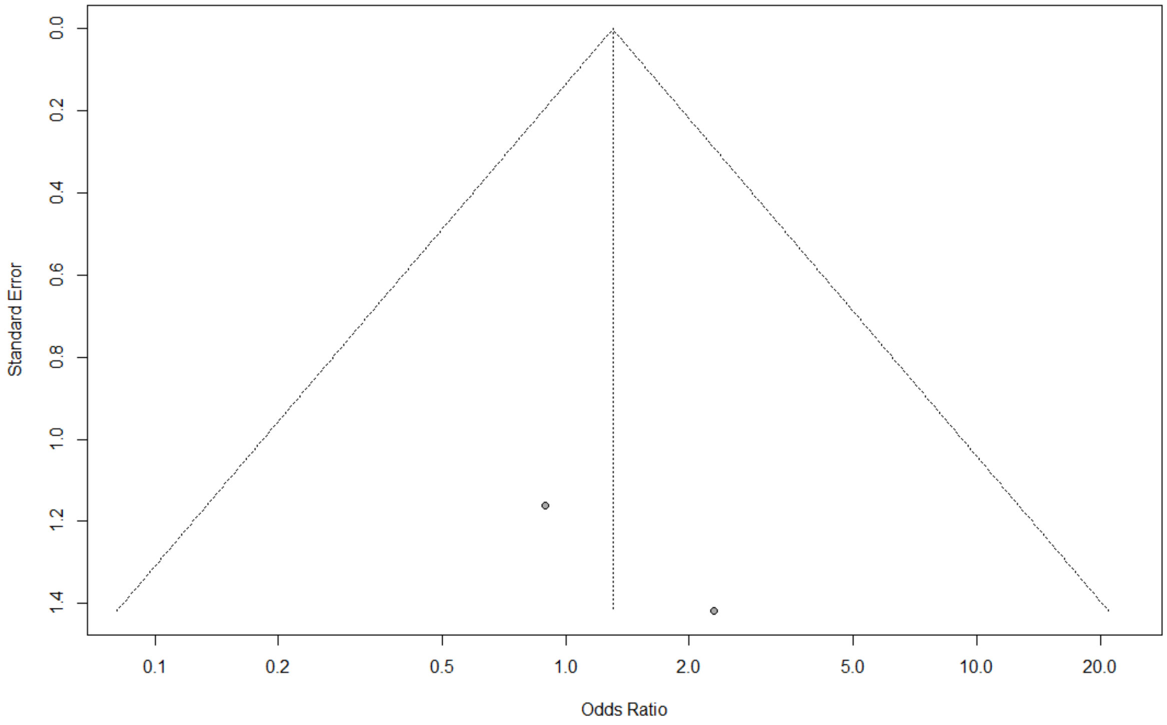

*Bleeding*

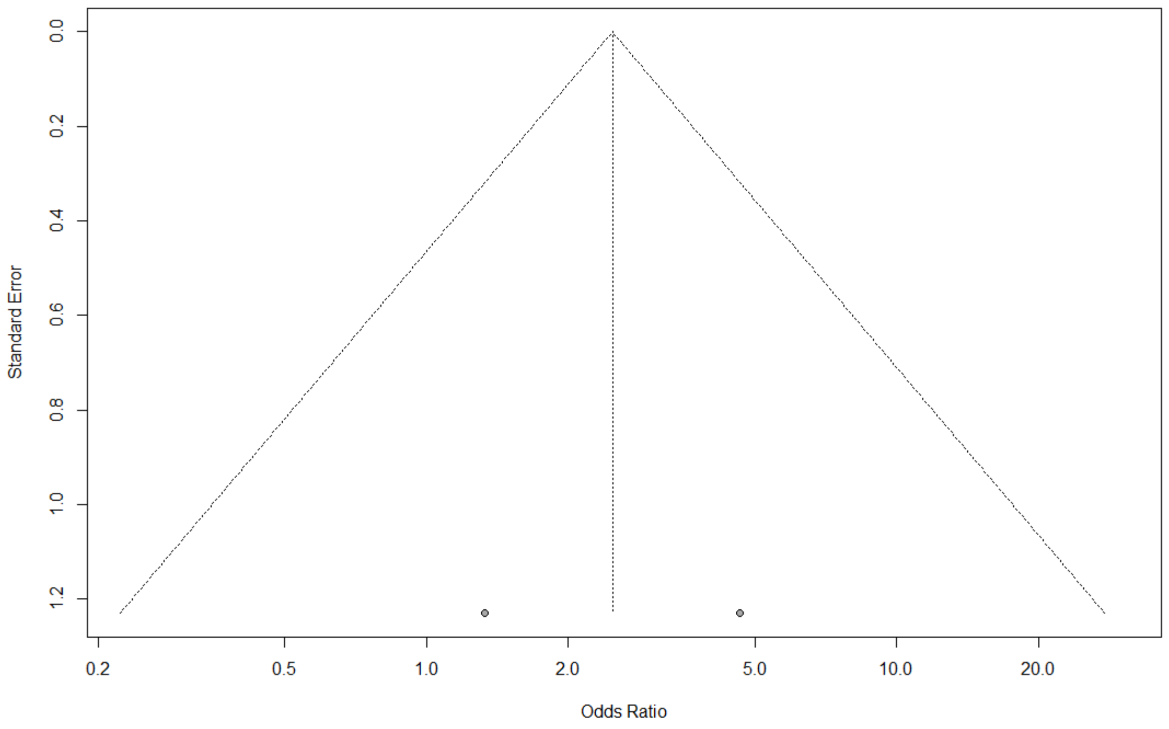

*Sepsis*

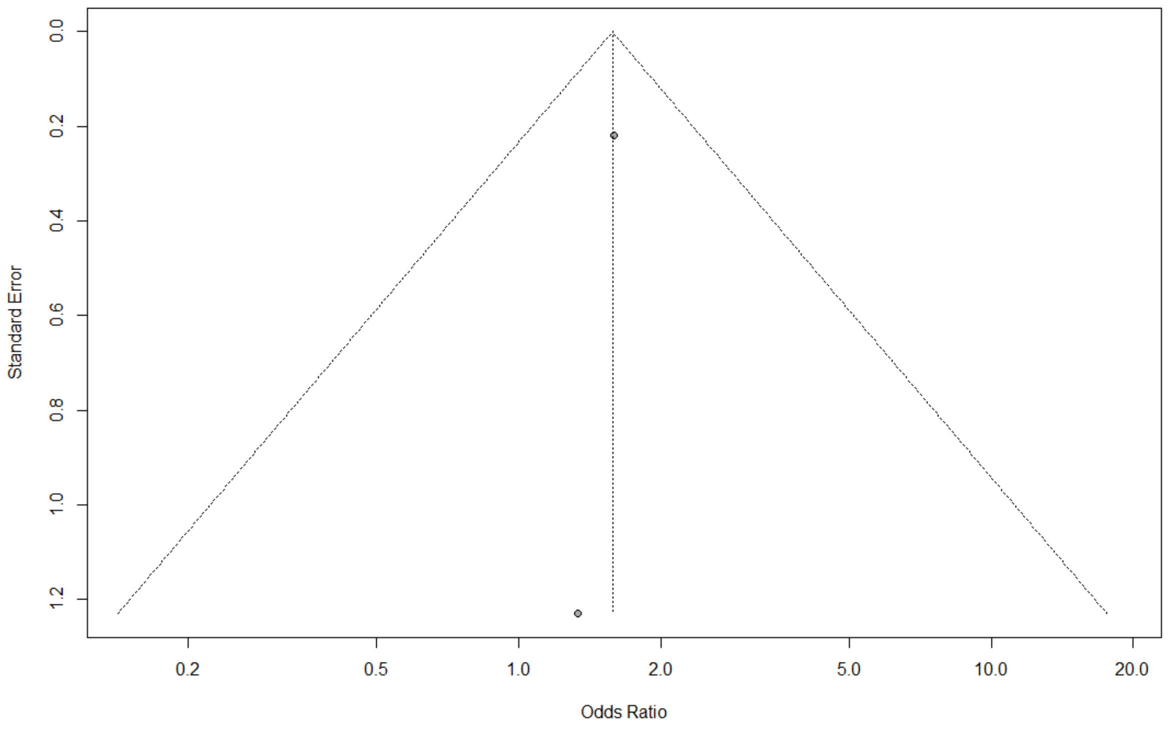

*Pneumonia*

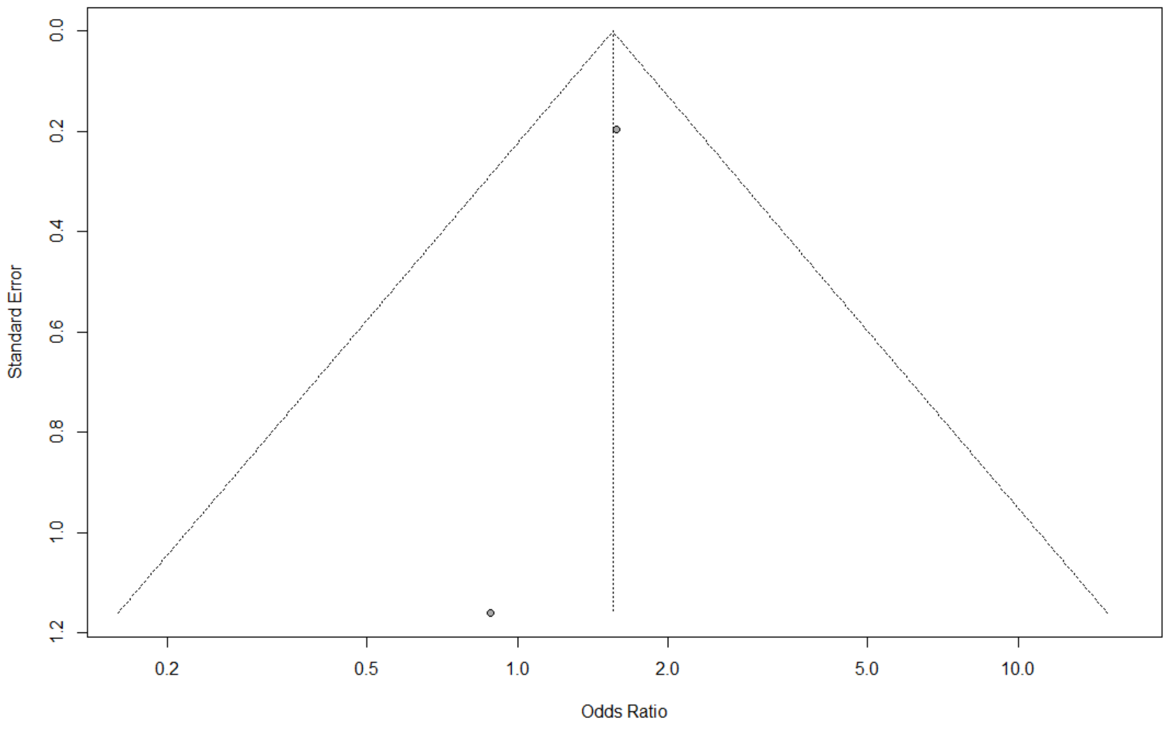

*__Readmission__*

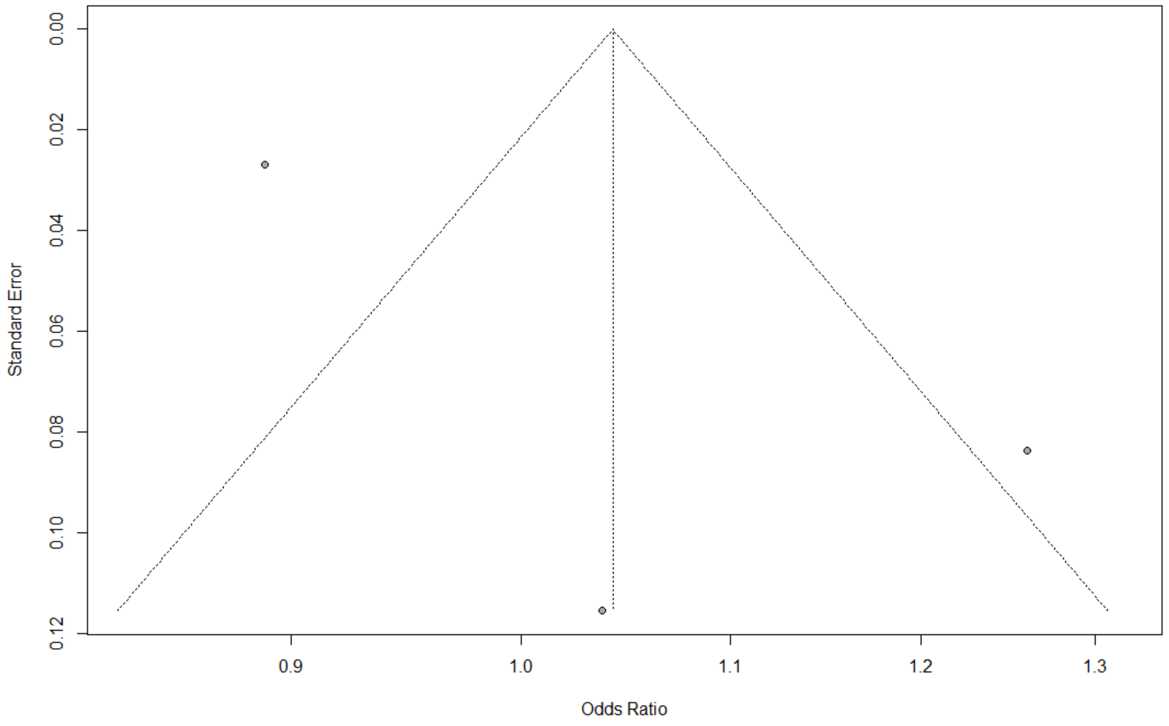

*__Mortality__*

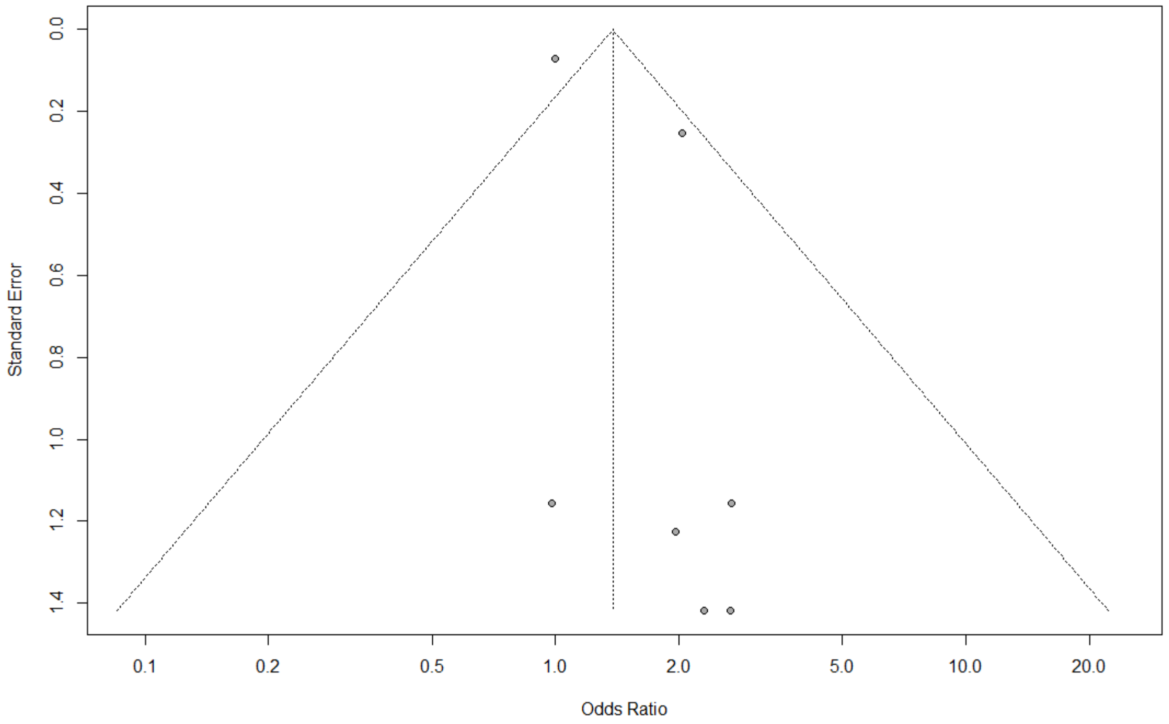

*Total length of stay*

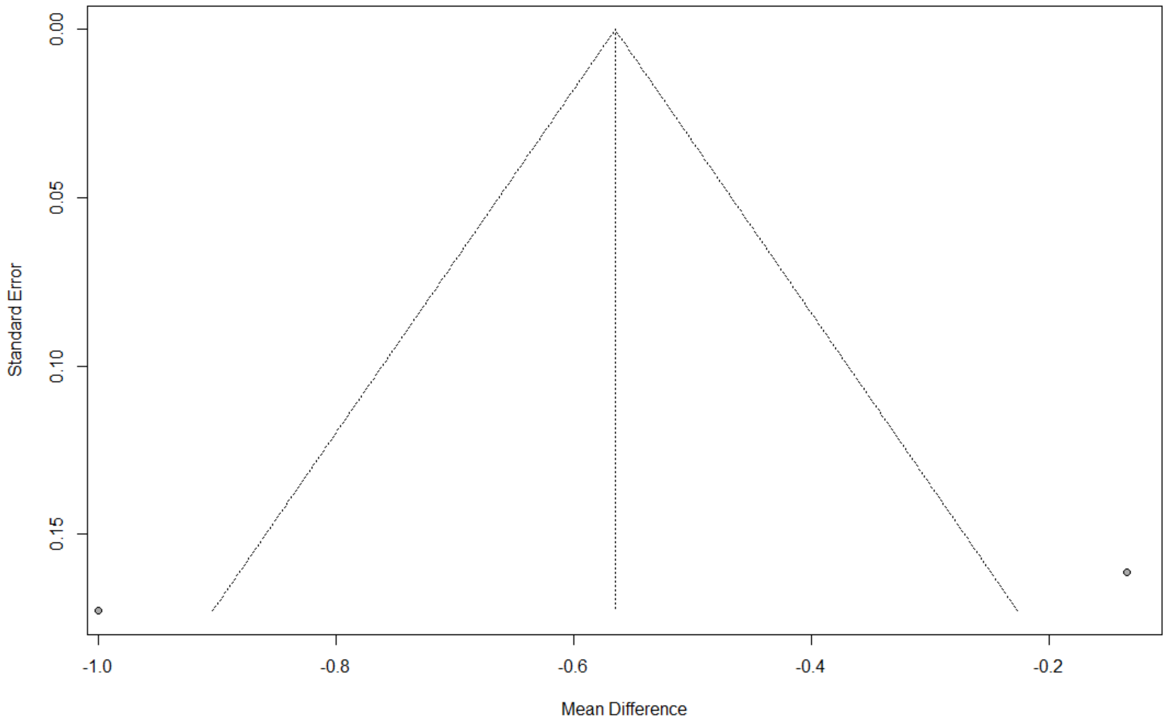
